# Epidemiology and costs of post-sepsis morbidity, nursing care dependency, and mortality in Germany

**DOI:** 10.1101/2021.02.25.21252347

**Authors:** Carolin Fleischmann-Struzek, Norman Rose, Antje Freytag, Melissa Spoden, Hallie C. Prescott, Anna Schettler, Lisa Wedekind, Bianka Ditscheid, Josephine Storch, Sebastian Born, Peter Schlattmann, Christian Günster, Konrad Reinhart, Christiane S. Hartog

## Abstract

**Purpose:** To quantify the frequency and co-occurrence of new diagnoses consistent with post-sepsis morbidity, mortality, new nursing care dependency, and total healthcare costs after sepsis.

**Methods:** Population-based cohort study using healthcare claims data from 23 million beneficiaries of a German health insurance provider. We included adult patients with incident hospital-treated sepsis identified by ICD-10 codes in 2013-2014. New medical, psychological and cognitive diagnoses associated with post-sepsis morbidity; mortality; dependency on nursing care; and total health care costs in survivors were assessed to 3 years after hospital discharge.

**Results:** Among 116,507 sepsis patients who survived hospitalization for sepsis, 74.3% had a new medical, psychological or cognitive diagnosis in the first year after discharge. 20.6% and 3.8% had new diagnoses in two and three domains, respectively. 31.5% were newly dependent on nursing care, and 30.7% died within the first year. In the second and third year, 65.8% and 59.4% of survivors had new diagnoses, respectively. Healthcare costs totaled an average 36,585 Euro/patient in three years, including index hospitalization costs. Occurrence of new diagnoses in predefined subgroups was: 73.7% (survivors of non-severe sepsis), 75.6% (severe sepsis), 78.3% (ICU-treated sepsis), 72.8% (non-ICU treated sepsis) and 68.5% (survivors without prior diagnoses).

**Conclusions:** New medical, psychological and cognitive diagnoses consistent with post-sepsis morbidity are common after sepsis, including among patients with less severe sepsis, no prior diagnoses, and younger age. This calls for more efforts to elucidate the underlying mechanisms, define optimal screening for common new diagnoses, and test interventions to prevent and treat post-sepsis morbidity.

**Trial Registration:** DRKS00016340

**Take home message:** This large population-based cohort of over 100,000 survivors of hospital-treated sepsis found high rates and a broad spectrum of new diagnoses consistent with post-sepsis morbidity, frequent new nursing care dependency, and high long-term mortality 1-3 years post sepsis. Post-sepsis morbidity was not limited to the oldest survivors or those with the most severe illness, but also affected younger survivors and those without pre-existing diagnoses.

## Introduction

Sepsis is a life-changing and disability-inducing event, resulting in considerable financial burden for healthcare systems [1-3]. An estimated 38 million patients survive sepsis each year [4], many of whom experience persisting health problems [1, 5], including new or worsened physical [6], psychological [7, 8] and cognitive [6] impairments. Because of these sequelae, sepsis survivors are often unable to return to work, need ongoing nursing care, and experience increased risk of death [9].

However, while the long-term consequences of sepsis are increasingly recognized, there are limited epidemiologic data on the co-occurrence of sepsis sequelae and the rate of sequelae in younger patients or those with less severe sepsis. In a nationwide US cohort of older sepsis survivors, one-sixth experienced persistent physical disability or cognitive impairment and one-third died during the following year [6, 10]. In ICU-treated sepsis survivors, there seems to be a considerable overlap with the “post-intensive care syndrome” (PICS) [11]. However, 50% of severe sepsis patients in the US [12] and two thirds of hospitalized sepsis patients in Germany [13] are not treated in an ICU. Data on the prevalence of sepsis sequelae in this patient group are lacking.

The present study used nationwide longitudinal claims data for one third of the German population to (i) quantify the frequency and co-occurrence of new medical, psychological, and cognitive diagnoses consistent with post-sepsis morbidity, (ii) compare the rate of mortality and new diagnoses by age groups, sepsis severity, ICU-treatment and pre-existing diseases and (iii) measure the cumulative costs of care.

## Methods

The study was pre-registered (DRKS00016340) and approved by the Jena University Hospital institutional review board (2019-1282-Daten).

### Data source

We performed a population-based observational cohort study using health claims data from the health insurer AOK (Allgemeine Ortskrankenkasse) from the years 2009-2017. Health insurance is mandatory in Germany; residents select any insurer and enroll without restriction. AOK is the largest, nationwide health insurer covering around 30% of the German population [14].

### Identification of sepsis patients

Patients ≥15 years with an inpatient hospitalization for sepsis (discharged 1/1/2013 through 12/31/2014) were identified by explicit ICD-10-GM-codes for sepsis (Supplement 1). We stratified the sepsis severity according to the sepsis-1/2 definitions [15, 16] as: sepsis – all forms; severe sepsis or septic shock (R65.1, R57.2); and non-severe sepsis. Coding of sepsis in Germany is rigorously controlled by the Medical Service of the Health Funds (MDK): Coding of non-severe sepsis in Germany is restricted to cases with positive blood culture and 2+ SIRS criteria or to cases with 4 SIRS criteria in case of negative blood cultures according to German coding regulations [17, 18]. The first hospital admission with sepsis during 2013-2014 was defined as the index hospitalization. We excluded patients with a prior diagnosis of sepsis occurring in the 2 years preceding hospitalization. Pre-existing diagnoses and comorbidities were assessed in a period up to five years prior to hospitalization. Therefore, patients who were not continuously insured by AOK from 01/01/2009 through the three year follow-up period the index hospitalization (or until death) were excluded.

### Characteristics of sepsis patients with index hospitalization

Patient demographics and clinical characteristics were assessed based on hospital discharge data, as well as a 12-month look-back in inpatient and outpatient claims. Study definitions are presented in Supplement 1. Prior nursing home residency and dependency on nursing care were determined based on graded care needs (which entitle patients to long-term care insurance benefits) [19].

### Determining new diagnoses and costs

New diagnoses consistent with post-sepsis morbidity were measured during the 1-12, 13-24 and 25-36 months after index hospital discharge. Based on a comprehensive literature review on post-sepsis morbidity, we identified relevant diagnoses and their associated ICD-10-GM codes (Supplement 1). Experts from intensive care, internal medicine, neurology, psychiatry, family medicine and rehabilitation medicine reviewed and approved the list of diagnoses consistent with post-sepsis morbidity. Diagnoses were grouped into three domains (medical, psychological, and cognitive). The medical domain included: respiratory, cardiovascular, cerebrovascular, renal, hepatic, metabolic, urogenital and neuromuscular/musculoskeletal diagnoses, sensory disorders, anaemia, fatigue, decubitus ulcer, pain, multidrug-resistant infections, complications of the tracheostomy and impairments of nutrition. The psychological domain included depression, anxiety, PTSD, sleeping disorders and substance abuse. The cognitive domain included mild to severe cognitive impairment as a single diagnosis. Both primary and secondary hospital discharge diagnoses as well as outpatient diagnoses labeled as confirmed diagnoses by the treating physicians [20] were included. We also assessed the co-occurrence of medical, cognitive, and psychological diagnoses; nursing care dependency; and post-discharge mortality. Finally, we used ICD-10-GM codes and OPS (Operationen- und Prozedurenschlüssel) codes to assess long-term mechanical ventilation and dialysis among sepsis survivors (Supplement 1).

We determined the prevalence of each diagnosis after sepsis; diagnoses were considered present if a relevant ICD-10 code was reported during at least one hospitalization or outpatient visit. Diagnoses were considered new if there was no related ICD-10 code during the 12-month look-back in inpatient and outpatient claims data. Survivors who did not have the particular diagnosis during the preceding 12 months were considered “at-risk” for incident diagnoses. Thus, incidence of each diagnosis in the 1^st^ year post sepsis was measured in only those patients at-risk (ie, did not have the diagnosis in the year prior to sepsis). Likewise, incident diagnoses in the 2^nd^ and 3^rd^ year survivors were measured among patients without the diagnosis through the 1^st^ and 2^nd^ years, respectively

We also measured total health care costs, including costs for hospitalizations, outpatient consultations, medications, treatments (*e.g.*, physical therapy, occupational therapy) and rehabilitation. In the German healthcare system, insured patients fulfilling certain requirements are entitled to treatment in a rehabilitation facility. There is no specific rehabilitation for sepsis. As data on outpatient diagnoses, cases, case-related therapies and corresponding costs in Germany are available on only a quarterly basis, data from quarters 1-4, 5-8 and 9-12 following the quarter of the index hospitalization discharge date were included in the 12, 24 and 36 month follow-up, respectively.

### Statistical Analyses

We reported continuous variables with means, standard deviations, medians and interquartile range (IQR) and categorical variables by proportions and 95% confidence intervals (CIs). Chi-square test and Welch-Satterthwaite t-tests were used for comparisons between subgroups (e.g. severe vs. non-severe sepsis, ICU-treated vs. non-ICU-treated). For all descriptive and inferential statistical analyses, SAS Enterprise Guide 7.1 was used. Kaplan-Meier estimates of the survivor functions were used for survival analyses. Differences in the survivor functions between subpopulations over follow-up were tested with the log-rank test. To facilitate the interpretation of survivor functions, non-parametric estimates of the hazard functions based on B-splines with 95% confidence bands are provided [21]. The software R was used for all survival analyses by means of the R packages ‘survival’ [22, 23] and ‘bshazard’[24].

## Results

Among 23.0 million eligible individuals, there were 159,684 index sepsis hospitalizations during 2013-2014 (353/100,000 person-years, Figure 1A). Sepsis patients were a mean age of 73.8 years and had modest comorbidity burden (mean unweighted Charlson Comorbidity Index 2.1). 38.3% were dependent on nursing care prior to sepsis hospitalization, and 11.7% resided in nursing homes prior to sepsis hospitalization. Only 6.7% had no pre-existing medical, psychological or cognitive diagnoses. 12.6% were employed prior to hospitalization.

**Figure 1:**
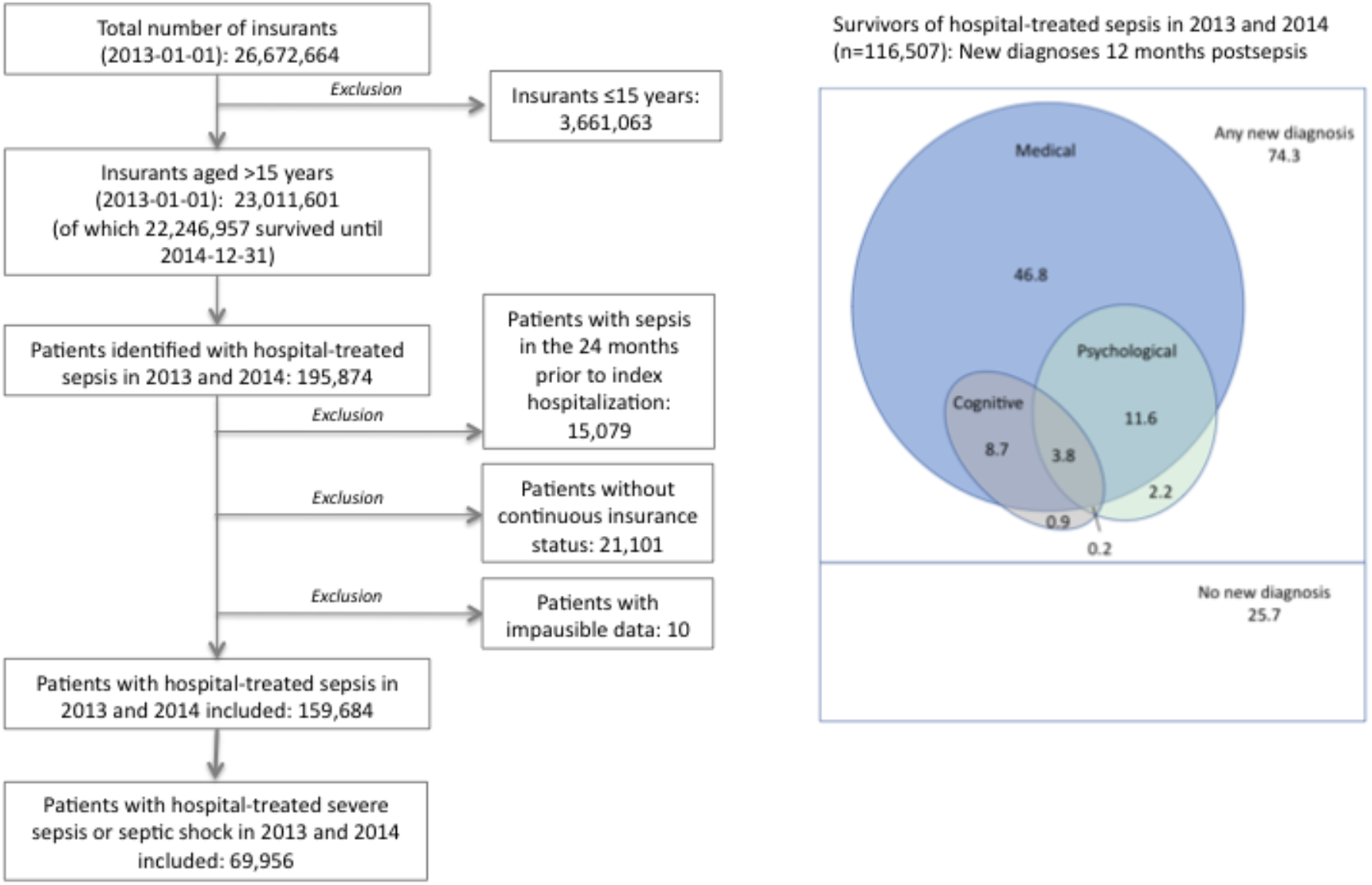
A) Flow of cohort, B) Postsepsis morbidity by domains and co-occurrence, in percent of survivors. This Euler. Diagram shows the proportion of survivors with new medical, cognitive, or psychological diagnoses in the first year. 74.3% had a new diagnosis in at least 1 domain, 20.6% had new diagnoses in at least 2 domains, and 25.7% had no new diagnosis.

Of 159,684 sepsis hospitalizations, 54,317 (34.0%) received intensive care. 89,728 (56.2%) had non-severe sepsis, while 69,956 (43.8%) had severe sepsis, including 20,589 (29.4%) with septic shock. Sepsis patients were hospitalized for a mean 20.6 days, and in-hospital mortality was 27.0% (Table 1). In-hospital mortality was higher in ICU-treated vs. non-ICU-treated patients (40.6% vs 20.0%, p<0.001), was higher for severe vs. non-severe sepsis (45.9% vs. 12.3%, p<0.001), and highest in septic shock (61.7%). Patients with no pre-existing medical, psychological or cognitive diagnoses had an in-hospital mortality of 19.2%. Overall, 5.5% of hospital survivors were discharged to inpatient rehabilitation. Detailed demographics and clinical features are reported in Supplement II Table S1.

**Table 1:**
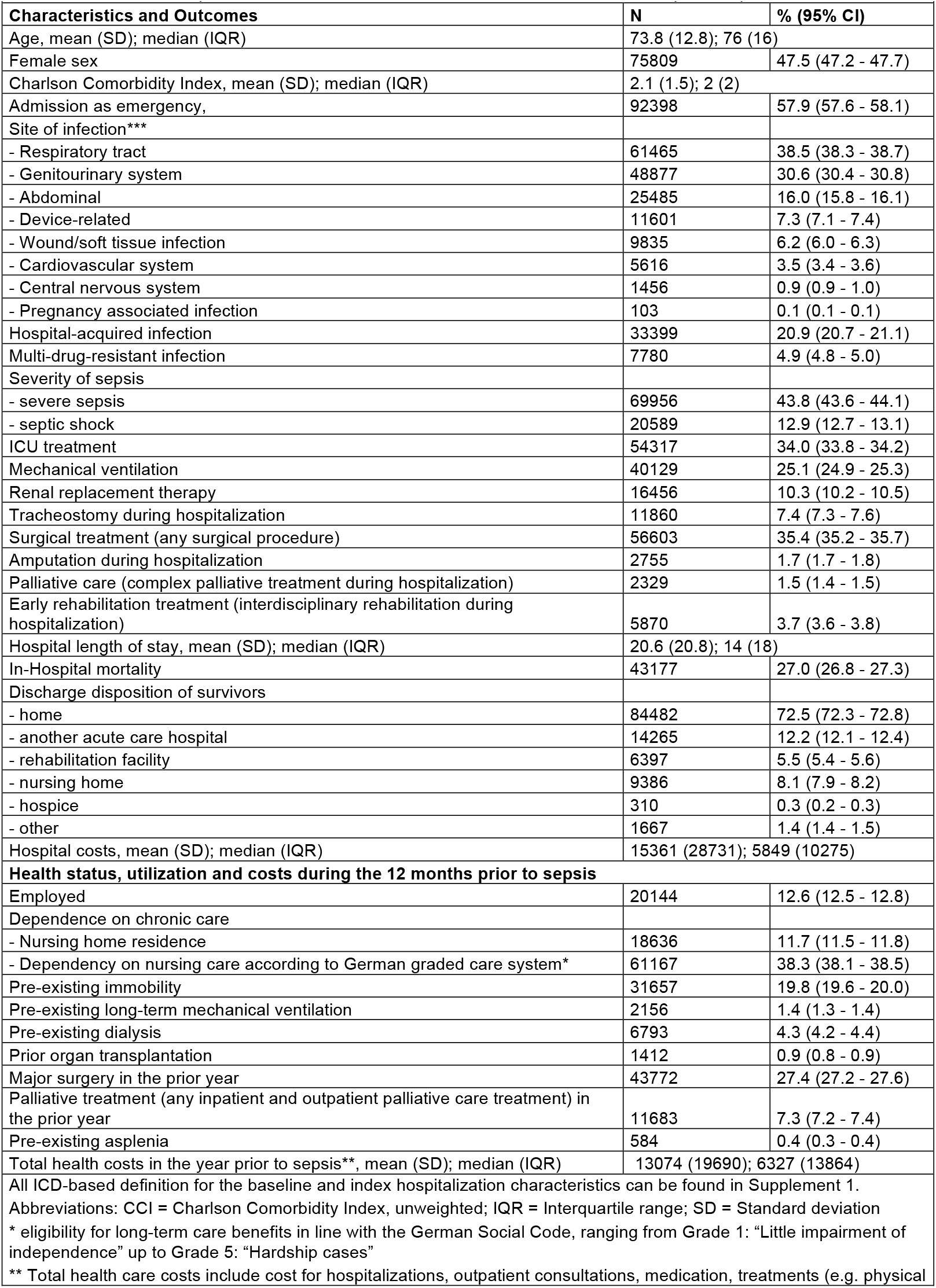
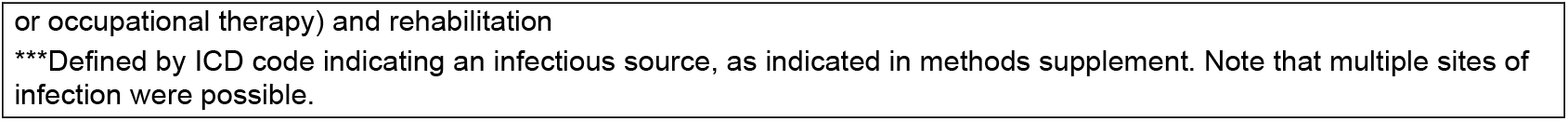
Patient and hospitalization characteristics and outcomes for 159,684 Index Sepsis Hospitalizations

|  |  |  |
| --- | --- | --- |
| Characteristics and Outcomes | N | % (95% CI) |
| Age, mean (SD); median (IQR) | 73.8 (12.8); 76 (16) |  |
| Female sex | 75809 | 47.5 (47.2 - 47.7) |
| Charlson Comorbidity Index, mean (SD); median (IQR) | 2.1 (1.5); 2 (2) |  |
| Admission as emergency, | 92398 | 57.9 (57.6 - 58.1) |
| Site of infection*** |  |  |
| - Respiratory tract | 61465 | 38.5 (38.3 - 38.7) |
| - Genitourinary system | 48877 | 30.6 (30.4 - 30.8) |
| - Abdominal | 25485 | 16.0 (15.8 - 16.1) |
| - Device-related | 11601 | 7.3 (7.1 - 7.4) |
| - Wound/soft tissue infection | 9835 | 6.2 (6.0 - 6.3) |
| - Cardiovascular system | 5616 | 3.5 (3.4 - 3.6) |
| - Central nervous system | 1456 | 0.9 (0.9 - 1.0) |
| - Pregnancy associated infection | 103 | 0.1 (0.1 - 0.1) |
| Hospital-acquired infection | 33399 | 20.9 (20.7 - 21.1) |
| Multi-drug-resistant infection | 7780 | 4.9 (4.8 - 5.0) |
| Severity of sepsis |  |  |
| - severe sepsis | 69956 | 43.8 (43.6 - 44.1) |
| - septic shock | 20589 | 12.9 (12.7 - 13.1) |
| ICU treatment | 54317 | 34.0 (33.8 - 34.2) |
| Mechanical ventilation | 40129 | 25.1 (24.9 - 25.3) |
| Renal replacement therapy | 16456 | 10.3 (10.2 - 10.5) |
| Tracheostomy during hospitalization | 11860 | 7.4 (7.3 - 7.6) |
| Surgical treatment (any surgical procedure) | 56603 | 35.4 (35.2 - 35.7) |
| Amputation during hospitalization | 2755 | 1.7 (1.7 - 1.8) |
| Palliative care (complex palliative treatment during hospitalization) | 2329 | 1.5 (1.4 - 1.5) |
| Early rehabilitation treatment (interdisciplinary rehabilitation during hospitalization) | 5870 | 3.7 (3.6 - 3.8) |
| Hospital length of stay, mean (SD); median (IQR) | 20.6 (20.8); 14 (18) |  |
| In-Hospital mortality | 43177 | 27.0 (26.8 - 27.3) |
| Discharge disposition of survivors |  |  |
| - home | 84482 | 72.5 (72.3 - 72.8) |
| - another acute care hospital | 14265 | 12.2 (12.1 - 12.4) |
| - rehabilitation facility | 6397 | 5.5 (5.4 - 5.6) |
| - nursing home | 9386 | 8.1 (7.9 - 8.2) |
| - hospice | 310 | 0.3 (0.2 - 0.3) |
| - other | 1667 | 1.4 (1.4 - 1.5) |
| Hospital costs, mean (SD); median (IQR) | 15361 (28731); 5849 (10275) |  |
| Health status, utilization and costs during the 12 months prior to sepsis |  |  |
| Employed | 20144 | 12.6 (12.5 - 12.8) |
| Dependence on chronic care |  |  |
| - Nursing home residence | 18636 | 11.7 (11.5 - 11.8) |
| - Dependency on nursing care according to German graded care system* | 61167 | 38.3 (38.1 - 38.5) |
| Pre-existing immobility | 31657 | 19.8 (19.6 - 20.0) |
| Pre-existing long-term mechanical ventilation | 2156 | 1.4 (1.3 - 1.4) |
| Pre-existing dialysis | 6793 | 4.3 (4.2 - 4.4) |
| Prior organ transplantation | 1412 | 0.9 (0.8 - 0.9) |
| Major surgery in the prior year | 43772 | 27.4 (27.2 - 27.6) |
| Palliative treatment (any inpatient and outpatient palliative care treatment) in the prior year | 11683 | 7.3 (7.2 - 7.4) |
| Pre-existing asplenia | 584 | 0.4 (0.3 - 0.4) |
| Total health costs in the year prior to sepsis**, mean (SD); median (IQR) | 13074 (19690); 6327 (13864) |  |
| All ICD-based definition for the baseline and index hospitalization characteristics can be found in Supplement 1. |  |  |
| Abbreviations: CCI = Charlson Comorbidity Index, unweighted; IQR = Interquartile range; SD = Standard deviation |  |  |
| * eligibility for long-term care benefits in line with the German Social Code, ranging from Grade 1: "Little impairment of independence" up to Grade 5: "Hardship cases" |  |  |
| ** Total health care costs include cost for hospitalizations, outpatient consultations, medication, treatments (e.g. physical |  |  |

or occupational therapy) and rehabilitation
\*\*\*Defined by ICD code indicating an infectious source, as indicated in methods supplement. Note that multiple sites of infection were possible.

Of the 116,507 patients who survived index hospitalization, 74.3% had a new medical, psychological, or cognitive diagnosis consistent with post-sepsis morbidity during their first year post-discharge (Figure 1B). Specifically, 70.9% had a new medical diagnosis, 17.9% had a new psychological diagnosis, and 18.5% of patients had a new cognitive diagnosis (Table 2). Among 74,878 hospital survivors without prior nursing care dependency, 31.5% had new nursing care dependency during the first year post-sepsis. 1.6% and 2.8% of at-risk survivors required new long-term mechanical ventilation and renal replacement therapy, respectively.

**Table 2:** Postsepsis diagnoses, mortality and costs over 3 years

| Table 2: Postsepsis diagnoses, mortality and costs over 3 years |  |  |  |  |  |  |
| --- | --- | --- | --- | --- | --- | --- |
|  | Timing of follow-up from index hospital discharge |  |  |  |  |  |
|  | 1-12 months |  | 13-24 months |  | 25-36 months |  |
| Outcomes among all survivors at start of the time period | N=116,507 | % (95% CI) | N=80,742 | % (95% CI) | N=68,940 | % (95% CI) |
| Survivors with any new diagnosis during the time period*, n, % of survivors | 86,578 | 74.3 (74.1 - 74.6) | 53,089 | 65.8 (65.4 - 66.1) | 40,959 | 59.4 (59.0 - 59.8) |
| New medical diagnosis, n, % of survivors | 82,629 | 70.9 (70.7 - 71.2) | 49,486 | 61.3 (61.0 - 61.6) | 3,7885 | 55.0 (54.6 - 55.3) |
| - Number of new medical diagnoses, mean (SD); median (IQR) | 2.2 (1.4); 2 (2) |  | 1.7 (1.0); 1 (2) |  | 1.7 (1.0); 1 (1) |  |
| New psychological diagnosis, n, % of survivors | 20,840 | 17.9 (17.7 - 18.1) | 10,296 | 12.8 (12.5 – 13.0) | 8,429 | 12.2 (12.0 - 12.5) |
| - Number of new medical diagnoses, mean (SD); median (IQR) | 2.2 (1.4); 2 (2) |  | 1.7 (1.0); 1 (2) |  | 1.7 (1.0); 1 (1) |  |
| New cognitive diseases, n/n at risk, % of survivors at risk | 15,955/86,350 | 18.5 (18.2 - 18.7) | 5,383/55,144 | 9.8 (9.5 – 10.) | 4,807/48,909 | 9.8 (9.6 - 10.1) |
| - Number of new medical diseases, mean (SD); median (IQR) | 2.2 (1.4); 2 (2) |  | 1.7 (1.0); 1 (2) |  | 1.7 (1.0); 1 (1) |  |
| New mechanical ventilation, n/n at risk, % of survivors at risk | 1,890/115,025 | 1.6 (1.6 - 1.7) | 906/78,999 | 1.1 (1.1 - 1.2) | 751/67,531 | 1.1 (1.0 - 1.2) |
| New dialysis, n/n at risk, % of survivors at risk | 3,144/111,993 | 2.8 (2.7 - 2.9) | 1,040/76,863 | 1.4 (1.3 - 1.4) | 789/65,925 | 1.2 (1.1 - 1.3) |
| New nursing home residence, n/n at risk, % survivors at risk | 12,485/103,912 | 12.0 (11.8 - 12.2) | 2,223/66,502 | 3.3 (3.2 - 3.5) | 1,950/57,409 | 3.4 (3.3 - 3.5) |
| New dependency on nursing care**, n/n at risk, % survivors at risk | 23,572/74,878 | 31.5 (31.1 - 31.8) | 3,784/40,925 | 9.2 (9.0 - 9.5) | 4,272/36,166 | 11.8 (11.5 - 12.1) |
| Mortality, n, % survivors | 35,765 | 30.7 (30.4-31.0) | 11,802 | 14.6 (14.4-14.9) | 9,082 | 13.2 (12.9 - 13.4) |
| Total health care costs [€]***, mean (SD); median (IQR) | 14,891 (24,737); 7,055 (14,958) |  | 11,503 (20,788); 5,040 (10,904) |  | 10,521 (19,146); 4,607 (9,803) |  |
| All ICD-based definition for the baseline and index hospitalization characteristics can be found in Supplement 1. |  |  |  |  |  |  |
| Abbreviations: CI = Confidence interval; IQR = Interquartile range; SD = Standard deviation |  |  |  |  |  |  |
| * at least one new cognitive, psychological or medical diagnosis in the respective time frame |  |  |  |  |  |  |
| ** eligibility for long-term care benefits in line with the German Social Code, ranging from Grade 1: “Little impairment of independence” up to Grade 5: “Hardship cases” |  |  |  |  |  |  |
| *** Total health care costs include cost for hospitalizations, outpatient consultations, medication and treatments (e.g. physical or occupational therapy) and rehabilitation |  |  |  |  |  |  |

The most common new diagnoses were neuromuscular/musculoskeletal, cardiovascular, respiratory, renal and urogenital diseases, occurring in 21% to 27% of at-risk survivors, respectively (Figure 2, Supplement II Table S2). New diagnoses of decubitus ulcers, chronic pain, and nutritional impairment occurred in 13-14% of at-risk survivors. New fatigue occurred in 8.2%, dysphagia in 6.9%, depression in 11.7 %, anxiety in 3.3%, and PTSD in 0.2%. 12.0% had new multi-drug resistant infections. New diagnoses in two and three domains occurred in 20.6% and 3.8% of survivors, respectively (Figure 1B). 39.9% had prevalent (pre-existing and new) diagnoses in two and 17.0% in all three domains in the first year post-sepsis.

**Figure 2:**
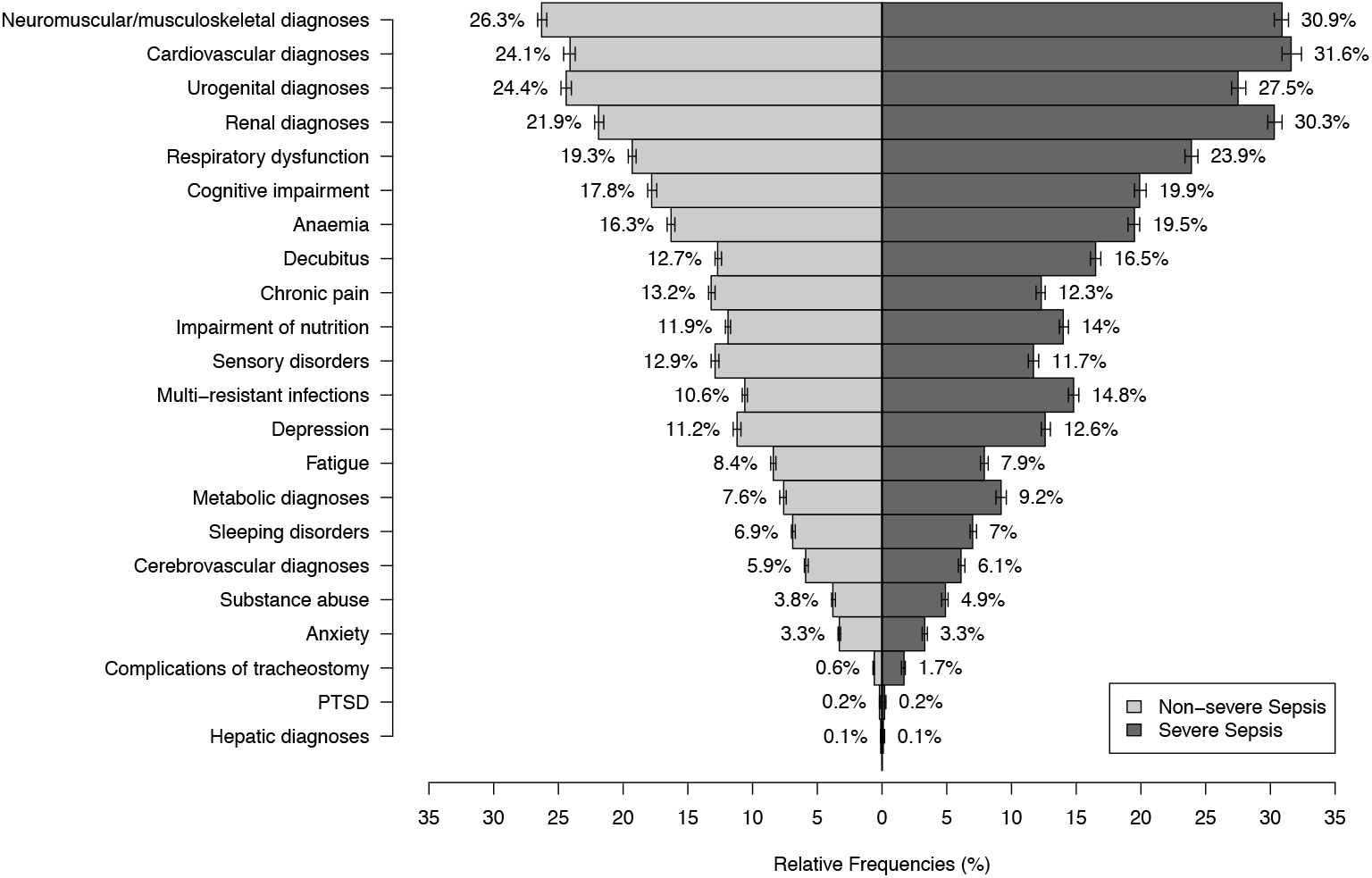
New postsepsis diagnoses in the 1-12 months after hospital discharge, in among survivors of non-severe vs. severe sepsis.

In the second and third year post-sepsis, new diagnoses occurred in 65.8% and 59.4% of one- and two-year survivors, respectively (Figure 3). While prevalence of cognitive diagnoses remained relatively stable, and prevalence of medical and psychological diagnoses increased, and the incidence of new diagnoses decreased across all domains among patients who survived over the one- and two-year period post-sepsis, respectively (Supplement II Table S3).

**Figure 3:**
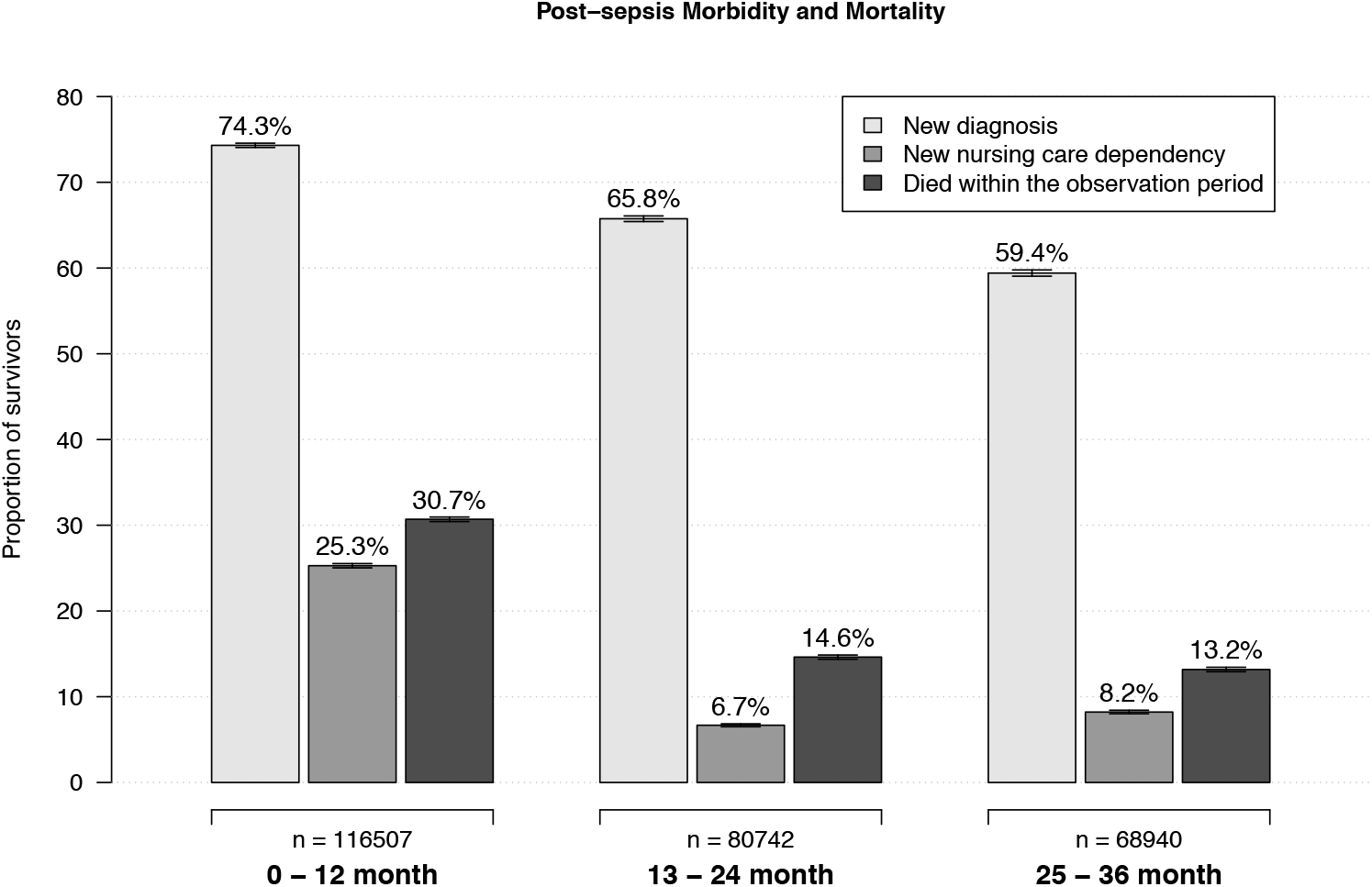
Postsepsis morbidity and mortality 1-12, 13-24 and 25-36 months after sepsis. **Legend** This figure shows the percentage of afflicted survivors among all sepsis survivors in the first, second and third year after sepsis. Of note, the proportion of patients with new nursing need is related to *all* sepsis survivors in this figure

New-onset diagnoses were more common among survivors of severe vs. non-severe sepsis (75.6% any new diagnosis, 72.8% new medical, 19.0% new psychological diagnosis, and 19.9% new cognitive), vs. 73.7% any new diagnosis, 70.0% new medical, 17.4% new psychological, and 17.8% new cognitive diagnosis), p<0.001 for each comparison, Table 3, Table 3A, Supplement II Table S4), and among ICU-treated vs. non-ICU treated sepsis (78.3% any new diagnosis, 75.3% new medical, 21.4% new psychological disease, and 20.2% new cognitive diagnosis vs. 72.8% any, 69.3% medical, 16.5% psychological and 17.8% cognitive), p<0.001 for each comparison, Table 3, Supplemental II Tables S5). Among survivors with no prior diagnoses, 63.5% had a new medical diagnosis, 25.0% had a new psychological diagnosis, and 12.8% had a new cognitive diagnosis (Table 3A, Supplement II Table S6, Supplement II Figure S1). Among survivors <40 years, 56.1% had at least one new diagnosis, and this proportion increased with greater age (Table 3B).

**Table 3A:**
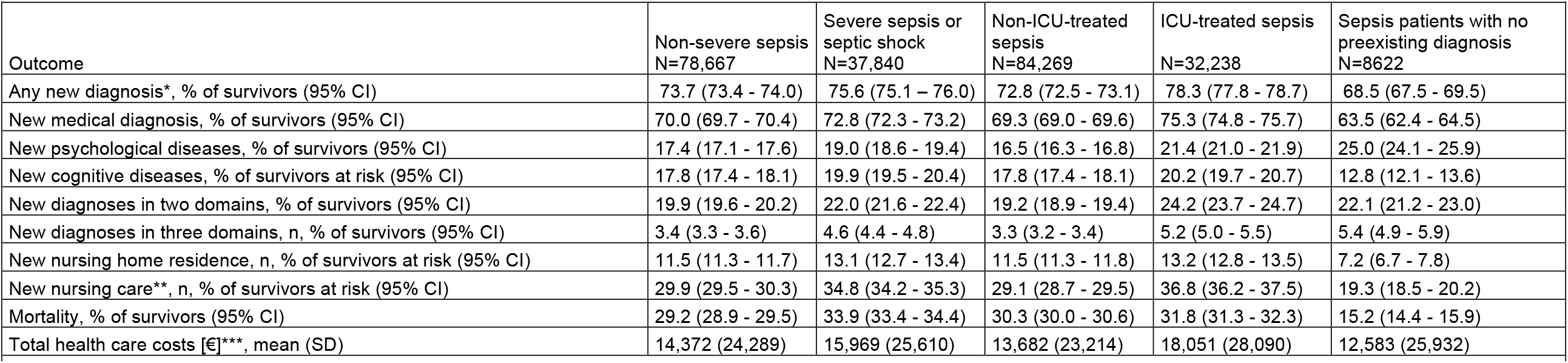
Postsepsis morbidity and mortality at 12 months, by patient subgroups

**Table 3B:**
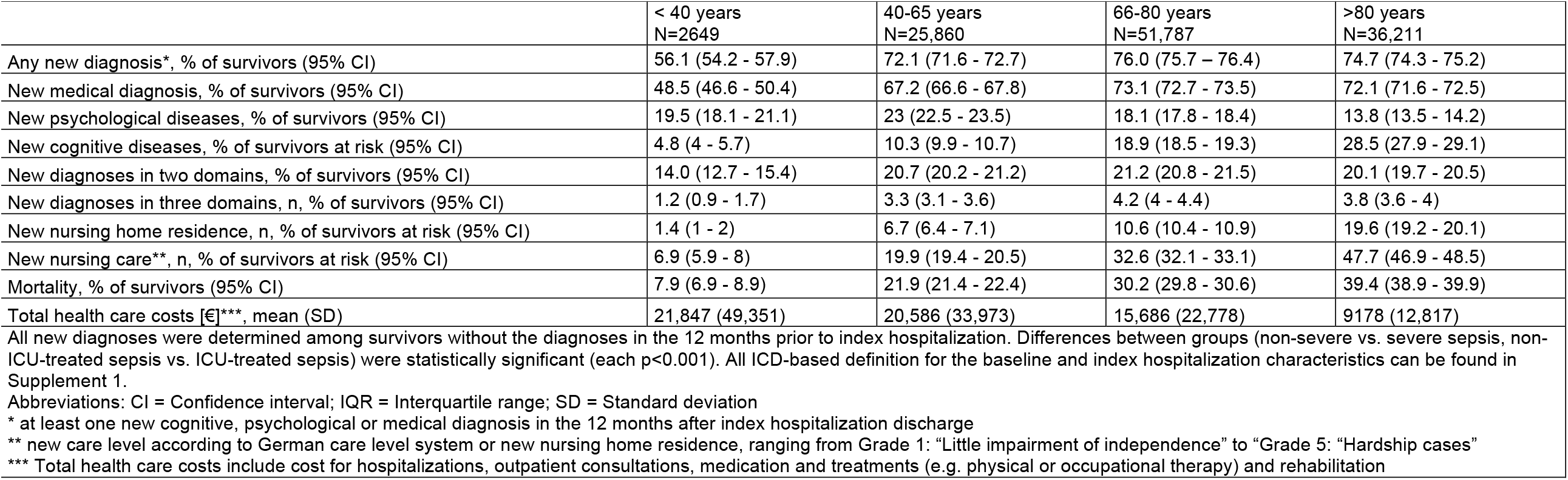
Postsepsis morbidity and mortality at 12 months, by age group

New nursing care was more common in survivors of severe vs. non-severe sepsis (34.8% vs. 29.9%, p<0.001), and in ICU-treated vs. non-ICU treated survivors (36.8% vs. 29.1%, p<0.001). 19.3% of survivors without pre-existing diseases required new nursing care. Among at-risk survivors aged >80 years, 47.7% required new nursing care. Likewise, new diagnoses in multiple domains were more common in severe vs. non-severe sepsis (22.0% vs. 19.9% with new diagnoses in two domains, p<0.001, and 4.6% vs. 3.4% in three domains, p<0.001), and in ICU-treated vs. non-ICU treated sepsis (24.2 vs. 19.2 and 5.2 vs. 3.3, respectively, each p<.001).

Among the 116,507 survivors of hospitalization for sepsis, one-year post-discharge mortality was 30.7%, and the majority of post-hospital deaths occurred within 100 days of hospital discharge (17.6% of hospitalization survivors died during the following 100 days, Figure 4A). One-year post-discharge mortality was higher in patients with severe vs non-sepsis sepsis, in ICU-treated vs non-ICU-treated sepsis, in patients with vs without pre-existing diagnoses (Figure 4B) and in older vs. younger patients. After approximately 100-150 days post-discharge, risk of subsequent mortality was similar between patients with severe vs. non-severe sepsis, and between ICU-treated vs. non-ICU-treated sepsis. However, mortality remained higher in older patients and patients with pre-existing diseases for the full 3 years of follow-up (Supplement II Figure S2a-e).

**Figure 4:**
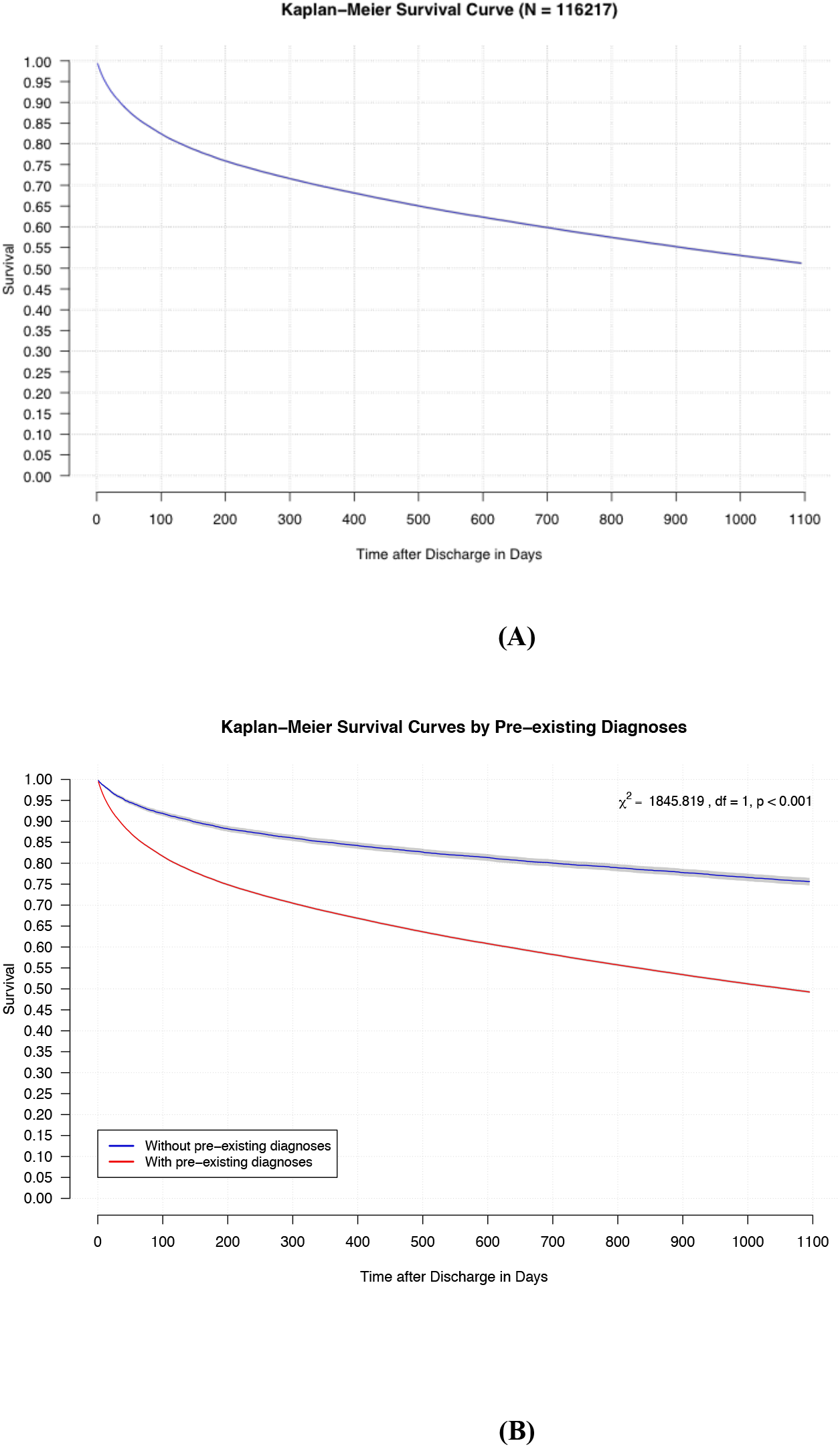
Kaplan-Meier Survival Curve of A) all sepsis patients, and B) sepsis patients according to pre-sepsis impairments, until 36 months after index hospital discharge. Survival analyses included all patients that survived the day of hospital discharge.

Among all sepsis survivors, mean health care costs were 14,891 Euro per patient in the first year and decreased to 11,503 Euro and 10,521 Euro in the second and third year, respectively (Table 2). For survivors of severe sepsis compared to survivors of non-severe sepsis, total health care costs were about 1,600 Euro higher in the first year (p<0.001). Similar differences were also seen in the following years (Supplement II Tab. S4). On average, the total costs for ICU-treated sepsis were about 4,400 Euro higher than for non-ICU-treated sepsis in the first year after sepsis (p<0.001). This difference was also seen – on a smaller extent – in the second and the third year (Supplement II Table S5).

Total health care costs from index hospitalization through the third year post-discharge were a mean of 36,585 Euro per patient (Supplement II Table S7). Total costs averaged 39,025 Euro for patients with severe sepsis, and 34,682 Euro for patients with non-severe sepsis.

## Discussion

In this large population-based cohort of over 100,000 survivors of hospital-treated sepsis with longitudinal follow-up to 3 years post-discharge, there were high rates of new diagnoses consistent with post-sepsis morbidity, new nursing care dependency, and death. Specifically, three-fourths of survivors were diagnosed with a new medical, psychological, or cognitive condition consistent with post-sepsis morbidity, and one-third died in the first year. Among survivors without pre-existing nursing care dependency, nearly one third were newly dependent on nursing care during the year following sepsis hospitalization. In the subsequent two years, more than half of survivors were diagnosed with a new condition, and one in six survivors died. New medical, psychological, and cognitive diagnoses co-occurred in a quarter of survivors. Importantly, and in contrast to many prior studies, this study captured a broad range of sepsis survivors, and showed that post-sepsis morbidity is not limited to the oldest survivors or those with the most severe illness—but also impacts younger survivors and those without pre-existing diagnoses.

With approximately 320,000 annual sepsis patients in Germany [13], the total direct costs for acute and three-year follow-up health care can be estimated at 11.7 billion Euro per year. The full economic impact of sepsis would be even higher if one considers reduced economic productivity of survivors, need for informal nursing care, and lifechanging effects on caregivers [25, 26], for whom many survivors depend on physical and financial support. These results highlight the considerable burden of sepsis and its long-lasting and multi-faceted sequelae for patients, families, and the healthcare system.

The rate of new diagnoses consistent with post-sepsis morbidity in our cohort may be higher than prior estimates. In a longitudinal cohort of older Americans, sepsis survivors acquired a median 1-2 new functional limitations, and 10.6% developed new moderate-severe cognitive impairment following sepsis [6]. This prior study suggested—based on the incidence of new functional and cognitive impairment—that sepsis was likely associated with substantial need for new nursing home placement and informal caregiving by family members, but was unable to measure these downstream impacts directly. By contrast, our study directly measured the incidence of new nursing care dependency and found that a third of at-risk sepsis survivors were newly dependent on nursing care, one-fifth had new cognitive diagnoses, and one-eight of at-risk survivors had a new diagnosis of depression.

Although the majority of survivors had new diagnoses consistent with post-sepsis morbidity, only 5.5% were discharged to rehabilitation facilities. Cardiovascular diseases were among the most common new diagnoses, likely with important mediator of long-term mortality in sepsis survivors [27]. The incidence of new pain diagnosis (13%) in our cohort is similar to a previous case-control study which found that 16% of ICU-treated sepsis and non-sepsis patients suffered from chronic pain at six months, but could detect no difference between the two groups [28]. Fatigue is a severely disabling symptom and an important determinant of quality of life for sepsis survivors [26]. Fatigue incidence in our study (8%) was in a similar range as reported by hospital-treated COVID-19 survivors [29]. This underscores that fatigue may also be associated with activation of the immune system [30]. Long-term ventilation is comparatively rare (1.6%) but nonetheless affects several thousand survivors yearly at enormous costs [31]. The incidence of anxiety (3.3%) or PTSD (0.2%) in our study was much lower than assessed among convenience samples of survivors from a sepsis self-help group (anxiety 60% and PTSD 69%) [32]. The difference can be explained because we assessed incident diseases in a population-based cohort. On the other hand, psychological diagnoses may be under-captured in our cohort, if physicians fail to provide a diagnosis for survivor symptoms [5].

Our study provides new insight into post-sepsis morbidity. We found that the majority of survivors are affected regardless of age, sepsis severity, ICU treatment, or pre-existing diseases. Furthermore, our findings demonstrate that new medical, psychological and cognitive diagnoses consistent with post-sepsis morbidity are also common among patients who fulfil the criteria of non-severe sepsis according to the old sepsis-1/2 criteria [15, 16].

These findings raise questions about the sensitivity of the new sepsis-3 definition in terms of the differentiation between patients with uncomplicated infections, which are less likely to cause major long-term morbidity, and patients with severe infections, formerly categorized as sepsis without organ dysfunction. SIRS criteria are no longer part of the current sepsis definition [33, 34], but the presence of at least two SIRS criteria in patients with proven or clinical suspected infection seems to identify an increased risk for major post-infection morbidity and hospital mortality.

Overall, our findings highlight the burden of post-sepsis morbidity and the need to develop and implement better systems to support survivors [35]. Our study has several strengths. First, to our knowledge, it is the first study to comprehensively investigate the epidemiology of post-sepsis morbidity across a population-based cohort of adult patients of all ages and with different severities of sepsis. The study used nationwide data of the largest healthcare insurance provider in Germany and covers approximately one-third of all German patients. Second, our study had a rigorous process to identify the specific diagnoses and diagnostic codes consistent with post-sepsis morbidity, based on a multi-professional panel of experts who care for patients after sepsis.

Several limitations need to be acknowledged. First, the identification of both sepsis patients and their subsequent diagnoses are dependent on the quality of coding. We used explicit sepsis codes for case identification, which have better specificity and positive predictive value than other strategies [36], but may miss some patients who meet clinical criteria for sepsis [12, 36]. Second, measuring post-sepsis morbidity based on diagnostic codes may result in misclassification. However, systematic screening of survivors for new medical, psychological, and cognitive diagnoses would not be feasible for such a large, population-based cohort. In Germany, the plausibility of inpatient and outpatient coding is audited by the Medical Services of the Health Care Funds and the Association of Statutory Health Insurance Physicians, which helps ensure the accuracy of coded diagnoses and mitigate the risk of misclassification in this study. Still, poor awareness of sepsis sequelae among patients and physicians may result in underdiagnosis. Third, our study is observational and cannot establish causality of post-sepsis morbidity. However, prior matched studies suggest that sepsis is causally associated with subsequent morbidity [9], particularly functional limitations, cognitive impairment, and select medical conditions. Fourth, our approach only allowed us to identify only new-onset diagnoses but not accelerated progression of pre-existing diagnoses. Fifth, our data lacked costs of emergency service utilization, transport, therapeutic aid prescriptions, dental care, homecare prescription, and nursing care, so underestimates the total financial toll of sepsis. Finally, we also cannot rule out that differences may exist in comparison with the general German patient population, but prior studies have suggested only small differences between AOK and non-AOK beneficiaries in Germany [37].

## Conclusion

In this large, population-based cohort of sepsis survivors with longitudinal follow-up to 3 years, we found that post-sepsis morbidity is common across all age groups and severities of sepsis. The financial toll of sepsis is substantial. This study underscores the need to raise awareness of post-sepsis morbidity, which is poorly known and understood by sepsis survivors, clinicians, and policy makers. Future research is needed to prevent, screen for, and treat post-sepsis morbidity. The development of comprehensive rehabilitation infrastructure also requires a better understanding of the mechanisms of long-term morbidity. finally, this study underpins the request of the World Health Assembly in its sepsis resolution, which asks member states to provide “access to relevant health care for survivors”.

## Data Availability

The authors confirm that the data utilized in this study cannot be made available in the manuscript, the supplemental files, or in a public repository due to German data protection laws (Bundesdatenschutzgesetz, BDSG). Therefore, they are stored on a secure drive in the AOK Research Institute (WIdO), to facilitate replication of the results. Generally, access to data of statutory health insurance funds for research purposes is possible only under the conditions defined in German Social Law (SGB V Paragraph 287). Requests for data access can be sent as a formal proposal specifying the recipient and purpose of the data transfer to the appropriate data protection agency. Access to the data used in this study can only be provided to external parties under the conditions of the cooperation contract of this research project and after written approval by the sickness fund. For assistance in obtaining access to the data, please contact.

## Acknowledgements

We thank Prof. Frank Oehmichen, Dr. Wolfgang Sauter, Dr. Konrad Schmidt, Prof. Winfried Meißner, PD Dr. Christoph Preul, PD Dr. Jenny Rosendahl, Dr. Romina Gawlytta and the SEPFROK expert panel (Prof. Horst Christian Vollmar, Prof. Uwe Janssens, Dr. Ruth Hecker, Arne Trumann, Dr. Simone Rosseau, Lothar Ullrich) for their contributions in the development of ICD-based definitions for postsepsis morbidity.

## Data availability statement

The authors confirm that the data utilized in this study cannot be made available in the manuscript, the supplemental files, or in a public repository due to German data protection laws (‘Bundesdatenschutzgesetz’, BDSG). Therefore, they are stored on a secure drive in the AOK Research Institute (WIdO), to facilitate replication of the results. Generally, access to data of statutory health insurance funds for research purposes is possible only under the conditions defined in German Social Law (SGB V § 287). Requests for data access can be sent as a formal proposal specifying the recipient and purpose of the data transfer to the appropriate data protection agency. Access to the data used in this study can only be provided to external parties under the conditions of the cooperation contract of this research project and after written approval by the sickness fund. For assistance in obtaining access to the data, please contact.

