## Supplement I for "Epidemiology and costs of post-sepsis morbidity, nursing care dependency, and mortality in Germany"

**Online Supplement I**

Postsepsis morbidity concept and operationalization	Page 2

Case identification: Definitions and Codes	Page 4

**Postsepsis morbidity concept and operationalization**

To identify diagnoses associated with postsepsis morbidity, we conducted a comprehensive literature review on reviews, round table/position papers, and large cohort studies investigating impairments following acute care treatment with sepsis or intensive care treatment.^1,9,11,36,39-41^ We classified diagnoses as medical, psychological, or cognitive (Table S1). For the identification of each diagnosis in hospital discharge and outpatient data, relevant ICD-10 codes or procedural codes were identified (i) from prior literature,^42-45^ or (ii) in the ICD-10-GM and list of procedural codes, to ensure completeness of definitions. The classification and ICD-10-GM definitions were reviewed in an iterative process by experts from the following fields:

- for sepsis rehabilitation Klinik Bavaria, Kreischa (Prof. Oehmichen, Dr. Sauter)
- from the SMOOTH^46^ study group (Dr. Konrad Schmidt, A. Freytag)
- from the REPAIR^47^ study group (PD Dr. Rosendahl, Dr. Gawlytta)
- for sepsis-related pain (Prof. Meißner) and neurology/geriatrics (PD Preul)
- from the SEPFROK expert panel (Prof. Vollmar (general medicine), Prof. Janssens (intensive care), Dr. Hecker (patient safety), A. Trumann (sepsis survivor), Dr. Rosseau (pulmonology), L. Ullrich (intensive care nursing)).

| **Table S1**: Definition of psychological, cognitive, and medical impairments following sepsis | | | |
| --- | --- | --- | --- |
|  | **Diagnosis associated with Postsepsis morbidity** | | |
| **Domains** | **Psychological** | **Cognitive** | **Medical** |
| **Diagnose** | PTSD | Cognitive dysfunction | Respiratory dysfunction |
|  | Depression |  | Cardiovascular disease  - Coronary heart disease and myocardial infarction  - Cardiomyopathy  - Heart failure  - Cardiac arrhythmias |
|  | Anxiety |  | Cerebrovascular disease |
|  | Sleeping disorders |  | Renal dysfunction |
|  | Substance abuse |  | Hepatic dysfunction |
|  |  |  | Metabolic diseases  - Diabetes mellitus  - Other metabolic diseases |
|  |  |  | Anaemia |
|  |  |  | Neuromuscular/musculoskeletal diseases  - ICUAW/CIP/CIM  - Dysphagia  - Voice disorders  - Contractures  - Immobility |
|  |  |  | Decubitus ulcer |
|  |  |  | Compl. of tracheostomy  - Tracheal stenoses |
|  |  |  | Urogenital diseases  - Incontinence  - Sexual disorders  - Urethral stricture |
|  |  |  | Sensory disorders  - Vestibular disorders  - Hearing disorder  - Taste and smelling disorders |
|  |  |  | Impairment of nutrition |
|  |  |  | Chronic pain |
|  |  |  | Infection with multi-resistant bacteria |
|  |  |  | Fatigue |

**Case identification: Definitions and Codes**

Sepsis

ICD-10-GM Codes: A02.1 - Salmonella sepsis, A20.0 - Bubonic plague, A20.7 - Septicaemic plague, A21.7 - Generalized tularaemia, A22.7 - Anthrax sepsis, A24.1 - Acute or fulminating melioidosis, A26.7 - Erysipelothrix sepsis, A28.2 - Extraintestinal yersiniosis, A32.7 - Listerial sepsis, A39.1 - Waterhouse-Friderichsen syndrome, A39.2-acute meningococcal sepsis, A39.3-chronic meningogoccal sepsis, A39.4 - Meningococcaemia, unspecified, A40.-Streptococcal sepsis, A41. - Other sepsis, A42.7 - Actinomycotic sepsis, A48.3 - Toxic shock syndrome, A49.9-Bacterial infection, unspecified, A54.8-Other gonococcal infections, B00.7 - Disseminated herpesviral disease, B37.6 - Candidal endocarditis, B37.7 - Candidal sepsis, B49 - Unspecified mycosis, O75.3 - Other infection during labour, O85 - other puerperal infections, R65.0 - Systemic Inflammatory Response Syndrome of infectious origin without organ failure, R65.1 - Systemic Inflammatory Response Syndrome of infectious origin with organ failure, R57.2 - Septic shock

Severe sepsis

ICD-10-GM Codes: R65.1 - Systemic Inflammatory Response Syndrome of infectious origin with organ failure

Septic shock

ICD-10-GM Codes: R57.2 - Septic shock

**Characteristics of sepsis and index treatment**

Assessed at discharge from the index treatment

Focus of infection

Respiratory tract

ICD-10-GM Codes: J01 - Acute sinusitis, J02 - Acute pharyngitis, J03 - Acute tonsillitis, J04 - Acute laryngitis and tracheitis, J06 - Acute upper respiratory infections of multiple and unspecified sites, J05 - Acute obstructive laryngitis [croup] and epiglottitis, J09 - Influenza due to identified zoonotic or pandemic influenza virus, J10 - Influenza due to identified seasonal influenza virus, J11 - Influenza, virus not identified, J12 - Viral pneumonia, not elsewhere classified, J13 - Pneumonia due to Streptococcus pneumoniae, J14 - Pneumonia due to Haemophilus influenzae, J15 - Bacterial pneumonia, not elsewhere classified, J16 - Pneumonia due to other infectious organisms, not elsewhere classified, J17 - Pneumonia in diseases classified elsewhere, J18 - Pneumonia, organism unspecified, J20 - Acute bronchitis, J21 - Acute bronchiolitis, J22 - Unspecified acute lower respiratory infection, J44.0 - Chronic obstructive pulmonary disease with acute lower respiratory infection, J44.1 - Chronic obstructive pulmonary disease with acute exacerbation, unspecified, J86 - Pyothorax, J85 - Abscess of lung and mediastinum, A15 - Respiratory tuberculosis, bacteriologicallyorhistologically confirmed, A16 - Respiratory tuberculosis, not confirmed bacteriologically or histologically, U69.00 - Hospital-acquired pneumonia in other diseases classified elsewhere, A36 - Diphtheria, A37 - Whooping cough, B38 - Coccidioidomycosis, B39 - Histoplasmosis

Abdominal infections

ICD-10-GM Codes: A00 - Cholera, A01 - Typhoid and paratyphoid fevers, A02 - Other salmonella infections, A03 - Shigellosis, A04 - Other bacterial intestinal infections, A05 - Other bacterial foodborne intoxications, not elsewhere classified, A06 - Amoebiasis, A07 - Other protozoal intestinal diseases, A08 - Viral and other specified intestinal infections, A09 - Other gastroenteritis and colitis of infectious and unspecified origin, K35 - Acute appendicitis, K37 - Unspecified appendicitis, K36 - Other appendicitis, K5702 - Diverticular disease of small intestine with perforation and abscess without bleeding, K5703 - Diverticular disease of small intestine with perforation and abscess with bleeding, K5712 - Diverticular disease of small intestine without perforation or abscess without bleeding, K57.13 - Diverticular disease of small intestine without perforation or abscess wit bleeding, K57.22 - Diverticular disease of large intestine with perforation and abscess without bleeding, K57.23 - Diverticular disease of large intestine with perforation, abscess and bleeding, K57.32 - Diverticular disease of large intestine without perforation or abscess wihout bleeding, K5733 - Diverticular disease of large intestine without perforation or abscess wit bleeding, K5742 - Diverticular disease of both small and large intestine with perforation and abscess without bleeding, K5743 - Diverticular disease of both small and large intestine with perforation, abscess and bleeding, K5752 - Diverticular disease of both small and large intestine without perforation or abscess or bleeding, K5753 - Diverticular disease of both small and large intestine without perforation or abscess with bleeding, K5782 - Diverticular disease of intestine, part unspecified, with perforation and abscess without bleeding, K5783 - Diverticular disease of intestine, part unspecified, with perforation, abscess and bleeding, K5792 - Diverticular disease of intestine, part unspecified, without perforation, abscess or bleeding, K5793 - Diverticular disease of intestine, part unspecified, without perforation or abscess with bleeding, K61 - Abscess of anal and rectal regions, K65 - Peritonitis, K67 - Disorders of peritoneum in infectious diseases classified elsewhere, K63.0 - Abscess of intestine, K63.1 - Perforation of intestine (nontraumatic), K75.0 - Abscess of liver, K75.1 - Phlebitis of portal vein, K81.0 - Cholecystitis, K77.0 - Liver disorders in infectious and parasitic diseases classified elsewhere, U69.40! - Recurrent infection due to Clostridium difficile

Wound/soft tissue infection

ICD-10-GM Codes: A46 - Erysipelas, B47 - Mycetoma, L03 - Phlegmon, L04 - Acute lymphadenitis, L08 - Other local infections of skin and subcutaneous tissue, L05 - Pilonidal cyst, B00 - Herpesviral [herpes simplex] infections, B07 - Viral warts, B08 - Other viral infections characterized by skin and mucous membrane lesions, not elsewhere classified, B09 - Unspecified viral infection characterized by skin and mucous membrane lesions, H05.0 - Acute inflammation of orbit, H60.2 - Malignant otitis externa, H70.0 - Acute mastoiditis, J36 - Peritonsillar abscess, J39.0 - Retropharyngeal and parapharyngeal abscess, J39.1 - Other abscess of pharynx, L02 - Cutaneous abscess, furuncle and carbuncle

Genitourinary system infection

ICD-10-GM Codes: N10 - Acute tubulo-interstitial nephritis, N15.1 - Renal and perinephric abscess, N15.9 - Renal tubulo-interstitial disease, unspecified, N34 - Urethritis and urethral syndrome, N30 - Cystitis, N39.0 - Urinary tract infection, site not specified, N41 - Inflammatory diseases of prostate, N45 - Orchitis and epididymitis, N48.2 - Other inflammatory disorders of penis, N49 - Inflammatory disorders of male genital organs, not elsewhere classified, N70 - Salpingitis and oophoritis, N71 - Inflammatory disease of uterus, except cervix, N72 - Inflammatory disease of cervix uteri, N73 - Other female pelvic inflammatory diseases, N74 - Female pelvic inflammatory disorders in diseases classified elsewhere, N75 - Diseases of Bartholin gland, N76 - Other inflammation of vagina and vulva, N77 - Vulvovaginal ulceration and inflammation in diseases classified elsewhere, N61 - Inflammatory disorders of breast, N98.0 - Infection associated with artificial insemination, A59 - Trichomoniasis, A55 - Chlamydial lymphogranuloma (venereum), A56 - Other sexually transmitted chlamydial diseases

Central nervous system infection

ICD-10-GM Codes: A39 - Meningococcal infection, G00 - Bacterial meningitis, not elsewhere classified, G01 - Meningitis in bacterial diseases classified elsewhere, G02 - Meningitis in other infectious and parasitic diseases classified elsewhere, G03 - Meningitis due to other and unspecified causes, G04 - Encephalitis, myelitis and encephalomyelitis, G05* - Encephalitis, myelitis and encephalomyelitis in diseases classified elsewhere, G06 - Intracranial and intraspinal abscess and granuloma, G07* - Intracranial and intraspinal abscess and granuloma in diseases classified elsewhere, G08 - Intracranial and intraspinal phlebitis and thrombophlebitis, A17+ - Tuberculosis of nervous system, A81 - Atypical virus infections of central nervous system, A83 - Mosquito-borne viral encephalitis, A84 - Tick-borne viral encephalitis, A85 - Other viral encephalitis, not elsewhere classified, A86 - Unspecified viral encephalitis, A87 - Viral meningitis, A88 - Other viral infections of central nervous system, not elsewhere classified, A89 - Unspecified viral infection of central nervous system

Cardiovascular system infection

ICD-10-GM Codes: I32 - Pericarditis in diseases classified elsewhere, I33 - Acute and subacute endocarditis, I39 - Endocarditis and heart valve disorders in diseases classified elsewhere, I40 - Acute myocarditis, I41 - Myocarditis in diseases classified elsewhere, I80 - Thombosis, phlebitis and thrombophlebitis, I38 - Endocarditis, valve unspecified, I98.1 - Cardiovascular disorders in other infectious and parasitic diseases classified elsewhere

Device-related infections

ICD-10-GM Codes: T82.6 - Infection and inflammatory reaction due to cardiac valve prosthesis, T82.7 - Infection and inflammatory reaction due to other cardiac and vascular devices , implants and grafts, T83.5 - Infection and inflammatory reaction due to prosthetic device, implant and graft in urinary system, T83.6 - Infection and inflammatory reaction due to prosthetic device, implant and graft in genital tract, T84.5 - Infection and inflammatory reaction due to internal joint prosthesis, T84.6 - Infection and inflammatory reaction due to internal fixation device [any site], T84.7 - Infection and inflammatory reaction due to other internal orthopaedic prosthetic devices, implants and grafts, T85.7 - Infection and inflammatory reaction due to other internal prosthetic devices, implants and grafts

Pregnancy associated infection

ICD-10-GM Codes: O75.3 - Other infection during labour, O85 - Puerperal fever, O030 - Spontaneous abortion; Incomplete, complicated by genital tract and pelvic infection, O035 - Spontaneous abortion; Complete or unspecified, complicated by genital tract and pelvic infection, O040 - Medical abortion; Incomplete, complicated by genital tract and pelvic infection, O045 - Medical abortion; Complete or unspecified, complicated by genital tract and pelvic infection, O050 - Other abortion; Incomplete, complicated by genital tract and pelvic infection, O055 - Other abortion; Complete or unspecified, complicated by genital tract and pelvic infection, O060 - unspecified abortion; Incomplete, complicated by genital tract and pelvic infection, O065 - Unspecified abortion; Complete or unspecified, complicated by genital tract and pelvic infection, O070--O07.5 - Failed medical abortion, complicated by genital tract and pelvic infection, O075 - Other and unspecified failed attempted abortion, complicated by genital tract and pelvic infection, O08.0 - Genital tract and pelvic infection following abortion and ectopic and molar pregnancy, O86 - Other puerperal infections, O23 - Infections of genitourinary tract in pregnancy, O41.1 - Infection of amniotic sac and membranes, O88.3 - Obstetric pyaemic and septic embolism, O91 - Infections of breast associated with childbirth, O98 - Maternal infectious and parasitic diseases classifiable elsewhere but complicating pregnancy, childbirth and the puerperium,

Hospital-acquired infections

ICD-10-GM Codes: T82.6 - Infection and inflammatory reaction due to cardiac valve prosthesis, T82.7 - Infection and inflammatory reaction due to other cardiac and vascular devices, implants and grafts, T84.5 - Infection and inflammatory reaction due to internal joint prosthesis, T84.6 - Infection and inflammatory reaction due to internal fixation device [any site], T84.7 - Infection and inflammatory reaction due to other internal orthopaedic prosthetic devices, implants and grafts, T85.72 - Infection and inflammatory reaction due to internal prosthetic devices, implants and grafts in the central nervous system, T85.73 - Infection and inflammatory reaction due to prosthetic devices or implants of the mamma, T85.75 - Infection and inflammatory reaction due to internal prosthetic devices, implants or grafts of the hepatobiliary system or pancreas, T85.76 - Infection and inflammatory reaction due to internal prosthetic devices, implants or grafts of the other gastrointestinal system, T85.78 - Infection and inflammatory reaction due to other internal prosthetic devices, implants and grafts, O86.0 - Infection of obstetric surgical wound, T83.5 - Infection and inflammatory reaction due to prosthetic device, implant and graft in urinary system, T83.6 - Infection and inflammatory reaction due to prosthetic device, implant and graft in genital tract, A04.7 - Enterocolitis due to Clostridium difficile, U69.40! - Recurrent infection due to Clostridium difficile, T80.2 - Infections following infusion, transfusion and therapeutic injection, T82.7 - Infection and inflammatory reaction due to other cardiac and vascular devices, implants and grafts, T81.4 - Infection following a procedure, not elsewhere classified, T85.71 - Infection and inflammatory reaction due to peritoneal dialysis catheter, T85.74 - Infection and inflammatory reaction due to percutaneous endoscopic gastrostomy/jejunostomy,T88.0 - Infection following immunization, U69.00 - Hospital-acquired pneumonia in patients aged 18 years or older

Multidrug-resistant infections

ICD-10-GM Codes: U80.! - Grampositive bacteria with specified antibiotic resistance, requiring special therapeutic or hygienic measures, U81.! - Gram - negative bacteria with specified antibiotic resistance, requiring special therapeutic or hygienic measures, U82.! - Mycobacteria with resistance against TB drugs (first line), U83.! - Candida with resistance against Fluconazole and Voriconazole, U84.! - Herpes virus with restistance against antivirals, U85! - Human Immunodeficiency Virus with resistance against antivirals or proteinase – inhibitors

OPS Codes: 8-987 - Complex treatment in the case of colonisation or infection with multidrug-resistant pathogens [MDR]

Organ dysfunction

Cardiovascular dysfunction/shock

ICD-10-GM Codes: I95.9 - Hypotension, unspecified, R57.8 - Other shock, R57.9 - Shock, unspecified, R57.2 - Septic shock

Respiratory dysfunction

ICD-10-GM Codes: J96. - Respiratory failure, not elsewhere classified, J80 - Adult respiratory distress syndrome, J98.4 - Other disorders of lung, R06.0 - Dyspnoea, R06.8 - Other and unspecified abnormalities of breathing

Encephalopathy

ICD-10-GM Codes: F05 - Delirium, not induced by alcohol and other psychoactive substances, G93.1 - Anoxic brain damage, not elsewhere classified, G93.4 - Encephalopathy, unspecified, R40 - Somnolence, stupor and coma

Renal dysfunction

ICD-10-GM Codes: N17. - Acute renal failure, N19 - Unspecified kidney failure,

Metabolic dysfunction

ICD-10-GM Codes: E87.2 - Acidosis

Abnormal coagulation

ICD-10-GM Codes: Coagulation D65 - Disseminated intravascular coagulation [defibrination syndrome], D68.8 - Other specified coagulation defects, D68.9 - Coagulation defect, unspecified, D69.5 - Secondary thrombocytopenia, D69.6 - Thrombocytopenia, unspecified,

Hepatic dysfunction

ICD-10-GM Codes: K72.0 Acute and subacute hepatic failure, K72.7 - Hepatic encephalopathy and hepatic coma, K72.9 - Hepatic failure, unspecified, K76.2 - Central haemorrhagic necrosis of liver, K76.3 - Infarction of liver,

Other organ dysfunction

ICD-10-GM Codes: R65.1 - Systemic Inflammatory Response Syndrome of infectious origin with organ complications

ICU Treatment

OPS Codes: 8-980 - Intensive care complex treatment, 8-98f - Costly intensive care complex treatment (basic procedure), 8-98d - Intensive care complex treatment in childhood (basic procedure), 8-98c - Intensive care complex treatment in childhood

Mechanical Ventilation

OPS Codes: 8-713 - Mechanical ventilation and respiratory support in adults, 8-712 - Mechanical ventilation and respiratory support in children and adolescents, 8-714 - Special procedure for mechanical ventilation in the case of severe respiratory failure, 870 - Access for mechanical ventilation and measures to maintain the airway, 871 - Mechanical ventilation and respiratory support via a mask or tube and ventilation weaning

Renal replacement therapy

OPS Codes: 8-853 - Haemofiltration, 8-854 - Haemodialysis, 8-855 - Haemodiafiltration, 8-857 - Peritoneal dialysis, 8-85a - Dialysis procedure due to a functional failure and failure of a kidney transplant

Tracheostomy during hospitalization

OPS Codes: 5-311 - Temporary tracheostomy, 5-312 - Permanent tracheostomy

Surgical treatment

OPS Codes: Any OPS Code from Chapter 5

Amputation during treatment

OPS Codes: 5-862 - Amputation and exarticulation of upper extremity, 5-863 - Amputation and exarticulation of hand, 5-864 - Amputation and exarticulation of lower extremity, 5-865 - Amputation and exarticulation of foot, 5-866 - Revision of amputation area

Palliative care

OPS Codes: 8-982 - Palliative medical complex treatment, 8-98e - Specialized inpatient palliative medical complex treatment, 898h - Specialized palliative medical complex treatment through a palliative care service

Early rehabilitation treatment

OPS Codes: 855 - Interdisciplinary and other early rehabilitation

Discharge disposition of survivors

| **Discharge disposition** | **Definition** |
| --- | --- |
| regular | regular termination of treatment, with or without post-discharge treatment intended |
| other hospital | transfer to another hospital;  transfer to another hospital as part of a cooperation;  external transfer for psychiatric treatment |
| hospice | discharge into a hospice |
| rehabilitation | discharge into a rehabilitation facility |
| nursing home | discharge into a long-term care facility |
| other | treatment terminated for other reasons, with or without post-discharge treatment intended;  Treatment terminated against medical advice, with or without post-discharge treatment intended;  Change of responsibility of the cost bearer;  Death;  internal routing;  Treatment terminated for other reasons, post-inpatient treatment  intended;  external transfer with relocation or change between the  Remuneration ranges of the DRG flat rate case, according to section 17b (1) first sentence of the Hospital Funding Act;  Internal transfer with a change between the DRG fee ranges  according to section 17b (1)first sentence of the Hospital Funding Act;  Relocation;  Discharge before resumption with reclassification;  Discharge before resumption with reclassification due to complication;  Discharge or transfer with subsequent readmission;  Case closure (internal transfer) when changing between full,  day-care and ward-equivalent treatment;  Start of an outside stay with an absence past midnight  (BPflV area - for the specialist department for laying);  Ending an outside stay with an absence past midnight  (BPflV area - for pseudo specialist department 0003);  Discharge at the end of the year if accepted in the previous year (for the purposes of  Billing - § 4 PEPPV);  Beginning of a period without direct patient contact  (station equivalent treatment);  Termination of a period without direct patient contact  (ward equivalent treatment - for pseudo-specialist department 0004); |

**12 months prior health and socioeconomic status**

Employment status

| **Employment Status** | **Definition** |
| --- | --- |
| Employed | Insurance type: 1 = compulsory health insurance, 2 = pension applicant, 5 = self-payer, 6 = rehabilitation |
| Unemployed | Type of insurance: 3 = pension recipient, 4 = benefits under the Employment Promotion Act, 9 = family insurance |

Comorbidities

defined according to Charlson Comorbidity Index^48^

Pre-existing immobility

ICD-10-GM Codes: R26.2 - Difficulty in walking, not elsewhere classified, R26.3 - Immobility, R29.6 - Tendency to fall, not elsewhere classified, Z99.3 - Dependence on wheelchair, Z74.0 - Need for assistance due to reduced mobility

Pre-existing long-term mechanical ventilation

ICD-10-GM Codes: Z99.0 - Dependence on aspirator, Z99.1 - Dependence on respirator

OPS Codes: 8-713 - Mechanical ventilation and respiratory support in adults, 8-712 - Mechanical ventilation and respiratory support in children and adolescents, 8-714 - Special procedure for mechanical ventilation in the case of severe respiratory failure, 870 - Access for mechanical ventilation and measures to maintain the airway, 871 - Mechanical ventilation and respiratory support via a mask or tube and ventilation weaning

Pre-existing dialysis

ICD-10-GM Codes: Z99.2 - Dependence on renal dialysis, Z49 - Care involving dialysis

OPS Codes: 5392 - Creation of an arteriovenous fistula, 8853 - Haemofiltration, 8854 - Haemodialysis, 8855 - Haemodiafiltration, 8857 - Peritoneal dialysis

Statutory scale of fees for physicians (GOÄ) Codes: 13602 - Flat rate supplementary fee for continuous care of a patient requiring dialysis, 13610 - Flat rate supplementary fee for medical care in the case of haemodialysis, peritoneal dialysis and special procedures, 13611 - Flat rate supplementary fee for medical care in the case of peritoneal dialysis, 04562 - Flat rate supplementary fee for continuous care of a patient requiring dialysis, 04564 - Flat rate supplementary fee for paediatric nephrology care when carrying out haemodialysis, 04565 - Flat rate supplementary fee for paediatric nephrology care when carrying out peritoneal dialysis, 40815 - Flat rate fee for dialysis in patients up to the age of 18 years at their place of residence, 40816 - Flat rate fee for peritoneal dialysis in patients up to the age of 18 years, 40817 - Flat rate fee for peritoneal dialysis in patients up to the age of 18 years at their place of residence, 40818 - Flat rate fee for haemodialysis in patients up to the age of 18 years during a holiday or other absence, 40819 - Flat rate fee for peritoneal dialysis in patients up to the age of 18 years during a holiday or other absence, 40823 - Flat rate fee for dialysis in insured persons from the age of 18 years, 40824 - Flat rate fee for dialysis in insured persons from the age of 18 years at their place of residence, 40825 - Flat rate fee for peritoneal dialysis in insured persons from the age of 18 years, 40826 - Flat rate fee for peritoneal dialysis in insured persons from the age of 18 years at their place of residence, 40827 - Flat rate fee for intermittent peritoneal dialysis in insured persons from the age of 18 years at their place of residence, 40828 - Flat rate fee for dialysis from the age of 18 years during a holiday or work-related stay, 40829 - Supplement to flat rate fee 40823 or 40825 for insured persons aged 59-69 years, 40830 - Supplement to flat rate fee 40824, 40826 and 40827 for insured persons aged 59-69 years, 40831 - Supplement to flat rate fee 40823 or 40825 for insured persons aged 69-79 years, 40832 - Supplement to flat rate fee 40824, 40826 and 40827 for insured persons aged 69-79 years, 40833 - Supplement to flat rate fee 40823 or 40825 for insured persons from 79 years of age, 40834 - Supplement to flat rate fee 40824, 40826 and 40827 for insured persons from 79 years of age, 40835 - Supplement to flat rate fee 40816, 40823 or 40825 for dialysis in a patient with an infection, 40836 - Supplement to flat rate fee 40815, 40817, 40818, 40819, 40824, 40826 to 40828 for dialysis in a patient with an infection, 40837 - Supplement to flat rate fee 40816 or 40825 for intermittent peritoneal dialysis, 40838 - Supplement to flat rate fee 40817, 40819, 40827 or 40828 for intermittent peritoneal dialysis

Prior organ transplantation

ICD-10-GM Codes: Z94 - Transplanted organ and tissue status

OPS Codes: 5504 - Liver transplantation, 5375 - Heart and heart-lung transplantation, 5555 - Kidney transplantation, 5335 - Lung transplantation, 55281 - Transplantation of a pancreas segment, 55282 - Transplantation of the pancreas (whole organ), 54676 - Small intestine transplantation

Prior major surgery

OPS Codes: 532 - Excision and resection of lung and bronchus, 533 - Other operations on lung and bronchus, 534 - Operations on the chest wall, pleura, mediastinum and diaphragm, 535 - Operations on the valves and septa of the heart and pericardial vessels, 536 - Operations on the coronary vessels, 537 - Surgical treatment of arrhythmias and other operations on the heart and pericardium, 8-851 - Bypass surgery (using the heart-lung machine), 538 - Incision, excision and occlusion of blood vessels., 539 - Other operations on blood vessels, 542 - Surgery on the oesophagus, 543 - Incision, excision and resection of the stomach, 544 - Extended stomach resection and other operations on the stomach, 545 - Incision, excision, resection and anastomosis of the small and large intestine, 546 - Other operations on the small and large intestine, 547 - Operations on the appendix, 548 - Operations on the rectum, 549 - Operations on the anus, 550 - Operations on the liver, 551 - Operations on the gallbladder and bile ducts, 552 - Operations on the pancreas, 553 - Abdominal hernia repair, 554 - Other operations in the abdominal region, 578 - Operations on other bones, 579 - Reduction of fractures and dislocations, 580 - Open joint surgery, 581 - Arthroscopic joint surgery, 582 - Prosthetic joint and bone replacement, 583 - Operations on the spine, 584 - Operations on the hand, 585 - Operations on muscles, tendons, fasciae and bursae, 586 - Replantation, exarticulation and amputation of extremities and other operations on the organs of locomotion, 501 - Incision (trepanation) and excision of the skull, brain and meninges, 502 - Other operations on the skull, brain and meninges, 503 - Operations on the spinal cord, spinal meninges and spinal canal, 504 - Operations on the nerves and nerve ganglia, 505 - Other operations on the nerves and nerve ganglia, 555 - Operations on the kidneys, 556 - Operations on the ureters, 557 - Operations on the bladder, 558 - Operations on the urethra, 559 - Other operations on the urinary organs, 560 - Operations on the prostate and seminal vesicles, 561 - Operations on the scrotum and tunica vaginalis testis, 562 - Operations on the testicles, 5-63 - Operations on the spermatic cord, epididymis and vas deferens, 564 - Operations on the penis, 565 - Operations on the ovary, 566 - Operations on the fallopian tubes, 567 - Operations for facial bone fractures, 568 - Incision, excision and removal of the uterus, 569 - Other operations on the uterus and operations on the parametria, 570 - Operations on the vagina and recto-uterine pouch, 571 - Operations on the vulva, 572 - Childbirth with breech presentation and instrumental delivery, 573 - Other operations to induce labour and during the birth, 574 - Caesarean section and child development, 575 - Other obstetric operations, 587 - Excision and resection of the breast, 588 - Other operations on the breast, 8-989 - Surgical complex treatment in cases of severe infection

Prior palliative treatment

ICD-10-GM Codes: Z51.5 - Palliative care

OPS: 8-982 - Palliative medical complex treatment, 8-98e - Specialized inpatient palliative medical complex treatment, 898h - Specialized palliative medical complex treatment through a palliative care service

Statutory scale of fees for physicians (GOÄ) Codes: 01425 - Initial care in specialized outpatient palliative care, 01426 - Follow-up prescription for continuation of the specialized outpatient palliative care, 03370 - Palliative medical initial diagnosis, 03371 - Supplementary fee for palliative medical care in the medical practice, 03372 - Supplementary fee for palliative medical care in the home, 03373 - Supplementary fee for palliative medical care in the home, 01425 - Initial prescription for specialized outpatient palliative care, 01426 - Follow-up prescription for continuation of the specialized outpatient palliative care, 03370 - Palliative medical initial diagnosis of patient status including treatment plan, 03371 - Supplementary fee to the insured persons flat rate 03000 for palliative medical care of the patient in the medical practice, 03372 - Supplementary fee to Catalogue of Tariffs for Physicians code 01410 or 01413 for palliative medical care in the home, 03373 - Supplementary fee to Catalogue of Tariffs for Physicians code 01411, 01412 or 01415 for palliative medical care in the home, 04370 - Palliative medical initial diagnosis, 04371 - Supplementary fee to the insured persons flat rate 04000 for palliative medical care of the patient in the medical practice, 04372 - Supplementary fee to Catalogue of Tariffs for Physicians code 01410 or 01413 for palliative medical care in the home, 04373 - Supplementary fee to Catalogue of Tariffs for Physicians code 01411, 01412 or 01415 for palliative medical care in the home, 37302 - Supplementary fee to the insured persons flat rate or basic flat rate for the coordinating panel doctor, 37314 - Consultation discussion doctor with an additional designation palliative medicine, 37318 - Telephone consultation, 37300 - Palliative medical initial diagnosis of patient status including treatment plan, 37305 - Supplementary fee to tariff codes 01410 and 01413 for palliative medical care in the home, 37306 - Supplementary fee to tariff codes 01411, 01412 and 01415 for palliative medical care in the home, 37317 - Supplementary fee to tariff code 37302 for accessibility and willingness to visit in critical phases, 37320 - Case conference

Pre-existing asplenia, coded in the five years prior to sepsis index hospitalization

ICD-10-GM Codes: Q89.0 - Asplenia (congenital), Q89.01 - Asplenia (congenital)

OPS Codes: 5-4131 - Splenectomy, total

**Postsepsis morbidity**

Cognitive impairment

ICD-10-GM Codes: F06.7 - Mild cognitive disorder, U51.- - Impairment of cognitive function, R41.0 - Disorientation, unspecified, F00* - Dementia in Alzheimer disease, F01 - Vascular dementia, F02* - Dementia in other diseases classified elsewhere, F03 - Unspecified dementia, F04 - Organic amnesic syndrome, not induced by alcohol and other psychoactive substances, F05 - Delirium, not induced by alcohol and other psychoactive substances, F06.9 - Unspecified organic mental disorder due to brain damage and dysfunction and to physical disease, F07.8 - Other organic personality and behavioural disorders due to brain disease, damage and dysfunction, F07.9 - Unspecified organic personality and behavioural disorder due to brain disease, damage and dysfunction, G30 - Alzheimer disease, G31.0 - Circumscribed brain atrophy, G31.1 - Senile degeneration of brain, not elsewhere classified, G31.9 - Degenerative disease of nervous system, unspecified, G32* - Other degenerative disorders of nervous system in diseases classified elsewhere

Psychological impairment

PTSD

ICD-10-GM Codes: F43 - Reaction to severe stress, and adjustment disorders, F43.0 - Acute stress reaction, F43.1 - Post-traumatic stress disorder, F43.2 - Adjustment disorders, F43.8 - Other reactions to severe stress, F43.9 - Reaction to severe stress, unspecified

Depression

ICD-10-GM Codes: F32 - Depressive episode, F33 - Recurrent depressive disorder, F34.1 - Dysthymia, F38 - Other mood [affective] disorders, F41.2 - Mixed anxiety and depressive disorder, F06.3 - Organic mood [affective] disorders

Anxiety

ICD-10-GM Codes: F40 - Phobic anxiety disorders, F41 - Other anxiety disorders, F06.4 - Organic anxiety disorder

Sleeping disorders

ICD-10-GM Codes: F51 - Nonorganic sleep disorders, G47 - Sleep disorders

Substance abuse

ICD-10-GM Codes: F10 - Mental and behavioural disorders due to use of alcohol, F11 - Mental and behavioural disorders due to use of opioids, F12 - Mental and behavioural disorders due to use of cannabinoids, F13 - Mental and behavioural disorders due to use of sedatives or hypnotics, F14 - Mental and behavioural disorders due to use of cocaine, F15 - Mental and behavioural disorders due to use of other stimulants, including caffeine, F16 - Mental and behavioural disorders due to use of hallucinogens, F17 - Mental and behavioural disorders due to use of tobacco, F18 - Mental and behavioural disorders due to use of volatile solvents, F19 - Mental and behavioural disorders due to multiple drug use and use of other psychoactive substances

Medical impairment

Respiratory dysfunction

ICD-10-GM Codes: J96 - Respiratory failure, not elsewhere classified, J98 - Other respiratory disorders, R06.0 - Dyspnoea, J80 - Adult respiratory distress syndrome

Cardiovascular diseases

Coronary heart disease and myocardial infarction

ICD-10-GM Codes: I20 - Angina pectoris, I21 - Acute myocardial infarction, I22 - Subsequent myocardial infarction, I24 - Other acute ischaemic heart diseases, I25 - Chronic ischaemic heart disease

Cardiomyopathy

ICD-10-GM Codes: I42 – Cardiomyopathy

Heart failure

ICD-10-GM Codes: I50 - Heart failure

Cardiac arrhythmias

ICD-10-GM Codes: I47 - Paroxysmal tachycardia, I48 - Atrial fibrillation and flutter, I49 - Other cardiac arrhythmias

Cerebrovascular diseases

ICD-10-GM Codes: I63 - Cerebral infarction, I64 - Stroke, not specified as haemorrhage or infarction, I65 - Occlusion and stenosis of precerebral arteries, not resulting in cerebral infarction, I66 - Occlusion and stenosis of cerebral arteries, not resulting in cerebral infarction

Renal diseases

ICD-10-GM Codes: N17 - Acute renal failure, N18 - Chronic kidney disease, N19 - Unspecified kidney failure

Hepatic diseases

ICD-10-GM Codes: K72.1 - Chronic hepatic failure

Metabolic diseases

Diabetes mellitus

ICD-10-GM Codes: E11 - Type 2 diabetes mellitus, E12 - Malnutrition-related diabetes mellitus, E13 - Other specified diabetes mellitus, E14 - Unspecified diabetes mellitus

Other metabolic diseases

ICD-10-GM Codes: E27 - Other disorders of adrenal gland, E35* - Disorders of endocrine glands in diseases classified elsewhere, E34.9 - Endocrine disorder, unspecified, E23 - Hypofunction and other disorders of pituitary gland

Anaemia

ICD-10-GM Codes: D50 - Iron deficiency anaemia, D51 - Vitamin B12 deficiency anaemia, D52 - Folate deficiency anaemia, D53 - Other nutritional anaemias, D63 - Anaemia in chronic diseases classified elsewhere, D64.9 - Anaemia, unspecified

Neuromuscular/musculoskeletal diseases

ICUAW/CIP/CIM

ICD-10-GM Codes: G62.8 - Critical illness polyneuropathy, G72.8 - Critical illness myopathy

Dysphagia

ICD-10-GM Codes: R13 - Dysphagia

Voice disorders

ICD-10-GM Codes: R49 - Voice disturbances

Contractures

ICD-10-GM Codes: M62.4 - Contracture of muscle, M24.5 - Contracture of joint, M25.6 - Stiffness of joint, not elsewhere classified, M21.62 - Acquired Pes equinus

Immobility

ICD-10-GM Codes: R26.2 - Difficulty in walking, not elsewhere classified, R26.3 - Immobility, R29.6 - Tendency to fall, not elsewhere classified, Z99.3 - Dependence on wheelchair, Z74.0 - Need for assistance due to reduced mobility

Decubitus

ICD-10-GM Codes: L89 - Decubitus ulcer and pressure area

Complications of tracheostomy

Complications of the tracheostoma

ICD-10-GM Codes: Z43.0 - Attention to tracheostomy, Z93.0 - Tracheostomy status, J95.0 - Tracheostomy malfunction

Tracheal stenoses

ICD-10-GM Codes: J95.5 - Postprocedural subglottic stenosis J95.81 - Tracheal stenosis following a procedure, J38.6 - Stenosis of larynx, J39.8 - Acquired tracheal stenosis

Urogenital diseases

Incontinence

ICD-10-GM Codes: R32 - Unspecified urinary incontinence, N39.3 - Stress incontinence, N39.4 - Other specified urinary incontinence, R15 - Faecal incontinence

Sexual disorders

ICD-10-GM Codes: F52 - Sexual dysfunction, not caused by organic disorder or disease

Urethral stricture

ICD-10-GM Codes: N99.1 - Postprocedural urethral stricture

Sensory disorders

Vestibular disorders

ICD-10-GM Codes: R42 - Dizziness and giddiness

Hearing disorder

ICD-10-GM Codes: H90 - Conductive and sensorineural hearing loss, H91 - Other hearing loss, H93 - Other disorders of ear, not elsewhere classified

Tase and smelling disorders

ICD-10-GM Codes: R43 - Disturbances of smell and taste

Impairment of nutrition

ICD-10-GM Codes: E41 - Nutritional marasmus, E43 - Unspecified severe protein-energy malnutrition, E44 - Protein-energy malnutrition of moderate and mild degree, E46 - Unspecified protein-energy malnutrition, R63.0 - Anorexia, R63.3 - Feeding difficulties and mismanagement, R63.4 - Abnormal weight loss, R63.6 - Insufficient intake of food and water, R63.8 - Other symptoms and signs concerning food and fluid intake, R64 - Cachexia

Multidrug-resistant infections

ICD-10-GM Codes: U80.-! - Gram-positive bacteria with specified antibiotic resistance, requiring special therapeutic or hygienic measures, U81.-! - Gram-negative bacteria with specified antibiotic resistance, requiring special therapeutic or hygienic measures, U82 - Mycobacteria with resistance against TB drugs (first line), U83 - Candida with resistance against Fluconazole and Voriconazole, U84 - Herpes virus with restistance against antivirals

Chronic pain

ICD-10-GM Codes: R52.1 - Chronic intractable pain, R52.2 - Other chronic pain, R52.9 - Pain, unspecified, F45.4 - Persistent somatoform pain disorder, F45.41 - Chronic pain disorder associated with psychological and behavioural factors, G54.6 - Phantom limb syndrome with pain

Fatigue

ICD-10-GM Codes: R53 - Malaise and fatigue, G93.3 - Chronic fatigue syndrome

Long-term mechanical ventilation

ICD-10-GM Codes: Z99.0 - Dependence on aspirator, Z99.1 - Dependence on respirator

OPS Codes: 8-713 - Mechanical ventilation and respiratory support in adults, 8-716 - Setting up of home mechanical ventilation, 8-718 - Weaning from mechanical ventilation

Dialysis

ICD-10-GM Codes: Z99.2 - Dependence on renal dialysis, Z49 - Care involving dialysis

OPS Codes: 8-853 - Haemofiltration, 8-854 - Haemodialysis, 8-855 - Haemodiafiltration, 8-857 - Peritoneal dialysis, 8-85a - Dialysis procedure due to a functional failure and failure of a kidney transplant
