## Supplement II for "Epidemiology and costs of post-sepsis morbidity, nursing care dependency, and mortality in Germany"

**Online Supplement II**

Table S1: Baseline characteristics of different patient groups	2

Table S2: Underlying diseases in the psychological and medical domain	4

Table S3: Prevalent and incident impairments in hospital survivors,
1-12, 13-24 and 25-36 months after sepsis	6

Table S4: Comparison of 1-12, 13-24 and 25-36 months outcomes and
costs of non-severe sepsis/severe sepsis survivors	7

Table S5: Comparison of 1-12, 13-24 and 25-36 months outcomes and
costs of ICU-treated/non-ICU-treated sepsis survivors	9

Table S6: 1-12, 13-24 and 25-36 months outcomes and costs of patients
without pre-existing impairments	10

Table S7: Costs	11

Fig. S1: Co-occurrence and mortality in patients 1-12 months after discharge
from the index hospitalization according to pre-existing impairments	12

Fig. S2: Hazard functions for death for a) all sepsis patients, b) severe and
non-severe sepsis patients, c) ICU- and non-ICU-treated sepsis patients, and
d) sepsis patients according to pre-existing impairments	14

| **Table S1:** Baseline characteristics of different patient groups | | | | | | | | | | | |
| --- | --- | --- | --- | --- | --- | --- | --- | --- | --- | --- | --- |
|  | **Non-severe sepsis** | | | **Severe sepsis** | | **ICU-treated sepsis** | | **Non-ICU-treated sepsis** | | **Sepsis patients w/o pre-existing impairments** | |
|  | **n** | **% (95% CI)** | | **n** | **% (95% CI)** | **n** | **% (95% CI)** | **n** | **% (95% CI)** | **n** | **% (95% CI)** |
| Patients, % of the total cohort | 89,728 | 56.2 (55.9 - 56.4) | | 69,956 | 43.8 (43.6 - 44.1) | 54,317 | 34 ( 33.8 - 34.2) | 105,367 | 66 (65.8 - 66.2) | 10,666 | 6.7 (6.6 - 6.8) |
| Age: mean (SD); median (IQR) | 73.8 (13.2); 76 (16) | | | 73.8 (12.3); 76 (16) | | 71.6 (12.3); 74 (16) | | 75 (12.9); 77 (14.8) | | 63.1 (16.3); 65.0 (22) | |
| Female sex | 44,246 | 49.3 (49 - 49.6) | | 31,563 | 45.1 (44.7 - 45.5) | 22,504 | 41.4 (41 - 41.8) | 53,305 | 50.6 (50.3 - 50.9) | 4,450 | 41.7 (40.8 - 42.7) |
| CCI: mean (SD); median (IQR) | 2 (1.4); 2 (2) | | | 2.3 (1.5); 2 (2) | | 2.4 (1.5); 2 (2) | | 2 (1.4); 2 (2) | | 1.2 (1.2); 1 (2) | |
| Admission as emergency | 51,208 | 57.5 (57.2 - 57.8) | | 41,185 | 59.2 (58.8 - 59.5) | 3,043 | 56.6 (56.2 - 57) | 61,650 | 58.5 (58.2 - 58.8) | 6,454 | 61(60 - 61.9) |
| Focus of infection, % | | | | | | | | | | | |
| - Respiratory tract | 27,561 | 30.7 (30.4 - 31) | | 33,904 | 48.5 (48.1 - 48.8) | 29,524 | 54.4 (53.9 - 54.8) | 31,941 | 30.3 (30 - 30.6) | 3,954 | 37.1 (36.2 - 38) |
| - Abdominal | 10,221 | 11.4 (11.2 - 11.6) | | 15,264 | 21.8 (21.5 - 22.1) | 13,215 | 24.3 (24 - 24.7) | 12,270 | 11.6 (11.5 - 11.8) | 1,967 | 18.4 (17.7 - 19.2) |
| - Wound/soft tissue infection | 5,305 | 5.9 (5.8 - 6.1) | | 4,530 | 6.5 (6.3 - 6.7) | 3,734 | 6.9 (6.7 - 7.1) | 6,101 | 5.8 (5.7 - 5.9) | 799 | 7.5 (7 - 8) |
| - Genitourinary system | 28,712 | 32 (31.7 - 32.3) | | 20,165 | 28.8 (28.5 - 29.2) | 14,952 | 27.5 (27.2 - 27.9) | 33,925 | 32.2 (31.9 - 32.5) | 2,756 | 25.8 (25 - 26.7) |
| - Central nervous system | 607 | 0.7 (0.6 - 0.7) | | 849 | 1.2 (1.1 - 1.3) | 882 | 1.6 (1.5 - 1.7) | 574 | 0.5 (0.5 - 0.6) | 212 | 2 (1.7 - 2.3) |
| - Cardiovascular system | 2,814 | 3.1 (3 - 3.3) | | 2,802 | 4 (3.9 - 4.2) | 2,529 | 4.7 (4.5 - 4.8) | 3,087 | 2.9 (2.8 - 3.0) | 439 | 4.1 (3.8 - 4.5) |
| - Device-related | 5,484 | 6.1 (6 - 6.3) | | 6,117 | 8.7 (8.5 - 9) | 6,455 | 11.9 (11.6 -12.2) | 5,146 | 4.9 (4.8 - 5.0) | 619 | 5.8 (5.4 - 6.3) |
| - Pregnancy associated infection | 84 | 0.1 (0.1 - 0.1) | | 19 | 0 (0 - 0) | 23 | 0 (0 - 0-1) | 80 | 0.1 (0.1 - 0.1) | 45 | 0.4 (0.3 - 0.6) |
| Hospital-acquired infection, % | 14,072 | 15.7 (15.4 - 15.9) | | 19,327 | 27.6 (27.3 - 28) | 19,089 | 35.1 (34.7 - 35.5) | 14,310 | 13.6 (13.4 - 13.8) | 1,986 | 18.6 (17.9 - 19.4) |
| Multi-resistant infection, % | 3,242 | 3.6 (3.5 - 3.7) | | 4,538 | 6.5 (6.3 - 6.7) | 4,623 | 8.5 (8.3 - 8.7) | 3,157 | 3 (2.9 - 3.1) | 312 | 2.9 (2.6 - 3.3) |
| Presence of any acute organ dysfunction, % | 40,175 | 44.8 (44.4 - 45.1) | | 62,329 | 89.1 (88.9 - 89.3) | 48,412 | 89.1 (88.9 - 89.4) | 54,092 | 51.3 (51 - 51.6) | 6,246 | 58.6(57.6 - 59.5) |
| number of organ dysfunctions: mean (SD); median (IQR) | 0.6 (0.8); 0 (1) | | | 2 (1.3); 2 (2) | | 2.1 (1.4); 2 (2) | | 0.8 (1); 1 (1) | | 1.2 (1.4); 2 (2) | |
| Occurance of septic shock, % | 0 | 0 (0-0) | | 20,589 | 29.4 (29.1 - 29.8) | 14,244 | 26.2 (25.9 - 26.6) | 6,345 | 6 (5.9 - 6.2) | 1,396 | 13.1 (12.5 - 13.7) |
| ICU treatment, % | 16,369 | 18.2 (18 - 18.5) | | 37,948 | 54.2 (53.9 - 54.6) | 54,317 | 100 (100-100) | 0 | 0 (0-0) | 4,096 | 38.4 (37.5 - 39.3) |
| Mechanical ventilation, % | 9,368 | 10.4 (10.2 - 10.6) | | 30,761 | 44 (43.6 - 44.3) | 32,086 | 59.1 (58.7 - 59.5) | 8,043 | 7.6 (7.5 - 7.8) | 3,037 | 28.5 (27.6 - 29.3) |
| Renal replacement therapy, % | 3,382 | 3.8 (3.6 - 3.9) | | 13,074 | 18.7 (18.4 - 19) | 12,893 | 23.7 (23.4 - 24.1) | 3,563 | 3.4 (3.3 - 3.5) | 937 | 8.8 (8.3 - 9.3) |
| Tracheostomy during hospitalization, % | 2,857 | 3.2 (3.1 - 3.3) | | 9,003 | 12.9 (12.6 - 13.1) | 10,636 | 19.6 (19.2 - 19.9) | 1,224 | 1.2 (1.1 - 1.2) | 1,158 | 10.9 (10.3 - 11.5) |
| Surgical treatment, % | 24,953 | 27.8 (27.5 - 28.1) | | 31,650 | 45.2 (44.9 - 45.6) | 33,093 | 69.9 (60.5 - 61.3) | 23,510 | 22.3 (22.1 - 22.6) | 4,664 | 43.7 (42.8 - 44.7) |
| Amputation during treatment, % | 1,168 | 1.3 (1.2 - 1.4) | | 1,587 | 2.3 (2.2 - 2.4) | 1,470 | 2.7 (2.6 - 2.8) | 1,285 | 1.2 (1.2 - 1.3) | 147 | 1.4 (1.2 - 1.6) |
| Palliative care, % | 1,454 | 1.6 (1.5 - 1.7) | | 875 | 1.3 (1.2 - 1.3) | 530 | 1.0 (0.9 (1.1) | 1,799 | 1.7 (1.6 - 1.8) | 98 | 0.9 (0.8 - 1.1) |
| Early rehabilitation treatment, % | 3,813 | 4.2 (4.1 - 4.4) | | 2,057 | 2.9 (2.8 - 3.1) | 1,644 | 3 (2.9 - 3.2) | 4,226 | 4 (3.9 - 4.1) | 204 | 1.9 (1.7 - 2.2) |
| Hospital length of stay: Mean, SD; Median, IQR, | 18.3 (17.7); 13 (14) | | | 23.4 (23.9); 16 (23) | | 30.4 (27.1); 23 (27) | | 15.5 (14.2); 11 (1) | | 21.8 (22.3); 14.0 (29) | |
| Hospital deaths, mortality, % | 11,061 | 12.3 (12.1 - 12.5) | | 32,116 | 45.9 (45.5 - 46.3) | 22,079 | 40.6 (40.2 - 41.1) | 21,098 | 20 (19.8 - 20.3) | 2,044 | 19.2 (18.4 - 19.9) |
| Discharge disposition of survivors, % | | | | | | | | | | | |
| - regular discharge | 60,940 | | 77.5 (77.2 - 77.7) | 23,542 | 62.2 (61.7 - 62.7) | 18,227 | 56.5 ( 56 - 57.1) | 66,255 | 78.6 (78.3 - 78.9) | 6,441 | 60.4 (59.5 - 61.3) |
| - other hospital | 7,079 | | 9 (8.7 - 9.3) | 7,186 | 19 (18.5 - 19.5) | 7,112 | 22.1 (21.6 - 22.5) | 7,153 | 8.5 (8.3 - 8.7) | 1,195 | 11.2 (10.3 - 12.2) |
| - rehabilitation | 3,092 | | 3.9 (3.6 - 4.2) | 3,305 | 8.7 (8.2 - 9.2) | 4,097 | 12.7 (12.3 - 13.1) | 2,300 | 2.7 (2.6 - 2.8) | 546 | 5.1 (4.2 - 6.1) |
| - nursing home | 6,120 | | 7.8 (7.5 - 8.1) | 3,266 | 8.6 (8.1 - 9.1) | 2,276 | 7.1 (6.8 - 7.3) | 7,110 | 8.4 (8.3 - 8.6) | 252 | 2.4 (1.4 - 3.3) |
| - hospice | 222 | | 0.3 (0 - 0.6) | 88 | 0.2 (0 - 0.7) | 86 | 0.3 (0.2 - 0.3) | 224 | 0.3 (0.2- 0.3) | 16 | 0.2 (0 - 1.1) |
| - other | 1,214 | | 1.5 (1.3 - 1.8) | 453 | 1.2 (0.7 - 1.7) | 440 | 1.4 (1.2 - 1.5) | 1,227 | 1.5 (1.4 - 1.5) | 2,216 | 20.8 (19.8 - 21.7) |
| Hospital costs: Mean, SD; Median (IQR) | 9,797 (18,446); 4,410 (5,205) | | | 22,496 (36,840); 9,281 (22,924) | | 31,656 (42,027); 17,587 (32,325) | | 6,960 (11,534); 3,928 (3,647) | | 18,232 (30,578); 6,086 (16,350) | |
| **Health status and health care use 12 months prior to index hospitalization** | | | | | | | | | | | |
| Employed persons, % | 11,865 | | 13.2 (13 - 13.4) | 8,279 | 11.8 (11.6 - 12.1) | 7,554 | 13.9 (13.6-14.2) | 12,590 | 11.9 (11.8 -12.1) | 3,638 | 34.1 (33.2 - 35) |
| Dependence on chronic care | | | | | | | | | | | |
| - Nursing home residence, % | 10,591 | | 11.8 (11.6 - 12) | 8,045 | 11.5 (11.3 - 11.7) | 4,058 | 4.5 (7.3 - 7.7) | 14,578 | 13.8 (13.6 - 14.0) | 3,638 | 34.1 (33.2 - 35) |
| - Care level per German care level system*, % | 34,364 | | 38.3 (38 - 38.6) | 26,803 | 38.3 (38 - 38.7) | 16,951 | 31.2 (30.8 - 31.6) | 44,216 | 42 (41.7 - 42.3) | 96 | 0.9 (0.7 - 1.1) |
| Pre-existing immobility, % | 17,490 | | 19.5 (19.2 - 19.8) | 14,167 | 20.3 (20 - 20.6) | 9,184 | 16.9 (16.6 - 17.2) | 22,473 | 21.3 (21.1 - 21.6) | 626 | 5.9 (5.4 - 6.3) |
| Pre-existing long-term mechanical ventilation, % | 1,024 | | 1.1 (1.1 - 1.2) | 1,132 | 1.6 (1.5 - 1.7) | 1,073 | 2.0 (1.9 - 2.1) | 1,083 | 1 (1-1.1) | 0 | 0 (0 - 0) |
| Pre-existing dialysis, % | 3,453 | | 3.8 (3.7 - 4) | 3,340 | 4.8 (4.6 - 4.9) | 2,865 | 5.3 (5.1 - 5.5) | 3,928 | 3.7 (3.6 - 3.8) | 0 | 0 (0 - 0) |
| Prior organ transplantation, % | 704 | | 0.8 (0.7 - 0.8) | 708 | 1 (0.9 - 1.1) | 580 | 1.1 (1.0 - 1.2) | 832 | 0.8 (0.7 - 0.8) | 3 | 0 (0 - 0.1) |
| Prior major surgery, % | 24,786 | | 27.6 (27.3 - 27.9) | 18,986 | 27.1 (26.8 - 27.5) | 14,791 | 27.2 (26.9 - 27.6) | 28,981 | 27.5 (27.2 - 27.8) | 25 | 0.2 (0.2 - 0.3) |
| Prior palliative treatment, % | 6,637 | | 7.4 (7.2 - 7.6) | 5,046 | 7.2 (7 - 7.4) | 2,945 | 5.4 (5.2 - 5.6) | 8,738 | 8.3 (8.1 - 8.5) | 992 | 9.3 (8.8 - 9.9) |
| Pre-existing asplenia, coded in the five years prior to sepsis index hospitalization, % | 343 | | 0.4 (0.3 - 0.4) | 241 | 0.3 (0.3 - 0.4) | 206 | 0.4 (0.3 - 0.4) | 378 | 0.4 (0.3 - 0.4) | 252 | 2.4 (2.1 - 2.7) |
| Total health care costs [€]**, mean (SD); median (IQR) | 12,982 (19,255); 6,194 (13,826) | | | 13,192 (20,233); 6,495 (13,912) | | 13,344 (21,175); 6,141 (14,003) | | 12,935 (18,877); 6,423 (13,792) | | 3,089 (9,001); 443 (1,799) | |
| IQR = Interquartile range; SD = Standard deviation  * eligibility for long-term care benefits in line with the German Social Code  ** Total health care costs include cost for hospitalizations, outpatient consultations, medication and treatments (e.g. physical or occupational therapy) and rehabilitation | | | | | | | | | | | |

| **Table S2: Underlying diseases in the psychological and medical domain** | | | | | | |
| --- | --- | --- | --- | --- | --- | --- |
|  | **Follow-Up after index admission** | | | | | |
|  | **12 months** | | **24 months** | | **36 months** | |
|  | **n** | **% (95 % CI)** | **n** | **% (95 % CI)** | **n** | **% (95 % CI)** |
| **Sepsis survivors, n** | **116,507** |  | **80,742** |  | **68,940** |  |
| **1. Medical Diagnoses** | | | | | | |
| ° Prevalence of physical diagnoses | | | | | | |
| °° respiratory dysfunction | 31,595 | 27.1 (26.9 - 27.4) | 19,514 | 24.2 (23.9 - 24.5) | 16,247 | 23.6 (23.3 - 23. 9) |
| °° cardiovascular diseases | 69,406 | 59.6 (59.3 - 59.9) | 49,877 | 61.8 (61.4 - 62.1) | 42,413 | 61.5 (61.2 - 61.9) |
| °°° coronary heart disease and myocardial infarction | 41,838 | 35.9 (35.6 - 36.2) | 30,724 | 38.1 (37.7 - 38.4) | 25,929 | 37.6 (37.3 - 38.0) |
| °°° cardiomyopathy | 4,949 | 4.2 (4.1 - 4.4) | 3,523 | 4.4 (4.2 - 4.5) | 3,004 | 4.4. (4.2 -4.5) |
| °°° heart failure | 43,178 | 37.1 (36.8 - 37.3) | 29,714 | 36.8 (36.5 - 37.1) | 25,423 | 36.9 (36.5 - 37.2) |
| °°° cardiac arrhythmias | 40,166 | 34.5 (34.2 - 34.7) | 28,349 | 35.1 (34.8 - 35.4) | 24,120 | 35.0 (34.6 - 35.3) |
| °° cerebrovascular diseases | 16,361 | 14.0 (13.8 - 14.2) | 12,016 | 14.9 (14.6 - 15.1) | 10,185 | 14.8 (14.5 - 15.0) |
| °° renal diseases | 49,446 | 42.4 (42.2 - 42.7) | 34,692 | 43.0 (42.6 - 43.3) | 30,068 | 43.6 (43.2 - 44) |
| °° hepatic diseases | 155 | 0.1 (0.1 - 0.2) | 80 | 01. (0.1 - 01.) | 59 | 01. (0.1 - 0.1) |
| °° metabolic diseases | 51,027 | 43.8 (43.5 - 44.1) | 37,984 | 47.0 (46.7 - 47.4) | 32,452 | 47.1 (46.7 - 47.4) |
| °°° diabetes mellitus | 50,369 | 43.2 (42.9 - 43.5) | 37,475 | 46.4 (46.1 - 46.8) | 32,033 | 46.5 (46.1 - 46.8) |
| °°° other metabolic diseases | 1,278 | 1.1 (1.0 - 1.2) | 978 | 1.2 (1.1 - 1.3) | 852 | 1.2 (1.2 - 1.3) |
| °° anaemia | 30,701 | 26.4 (26.1 - 26.6) | 19,044 | 23.6 (23.3 - 23.9) | 15,913 | 23.1 (22.8 - 23.4) |
| °° neuromuscular/musculoskeletal diseases | 40,962 | 35.2 (34.9 - 35.4) | 27,069 | 33.5 (33.2 - 33.9) | 23,333 | 33.8 (33.5 - 34.2) |
| °°° ICUAW/CIP/CIM | 2,659 | 2.3 (2.2 -2.4) | 1,334 | 1.7 (1.6 -1.7) | 1,103 | 1.6 (1.5 - 1.7) |
| °°° dysphagia | 10,285 | 8.8 (8.7 - 9.0) | 5,751 | 7.1 (6.9 - 7.3) | 4,688 | 6.8 (6.6 - 7.0) |
| °°° voice disorders | 1,337 | 1.1 (1.1 -1.2) | 956 | 1.2 (1.1 - 1.3) | 818 | 1.2 (1.1 - 1.3) |
| °°° contractures | 2,732 | 2.3 (2.3 - 2.4) | 1,893 | 2.3 (2.2 - 2.5) | 1,508 | 2.2. (2.1 - 2.3) |
| °°° immobility | 33,707 | 28.9 (28.7 - 29.2) | 22,594 | 28.0 (27.7 - 28.3) | 19.688 | 28.6 (28.2 - 28.9) |
| °° decubitus | 19,774 | 17.0 (16.8 - 17.2) | 9,839 | 12.2 (12.0 - 12.4) | 7,885 | 11.4 (11.2 - 11.7) |
| °° complications of the tracheostoma | 954 | 0.8 (0.8 - 0.9) | 385 | 0.5 (0.4 - 0.5) | 222 | 0.3 (0.3 - 0.4) |
| °°° tracheal stenoses | 448 | 0.4( 0.4 - 0.4) | 275 | 0.3 (0.3 -0.4) | 186 | 0.3 (0.2 - 0.3) |
| °° urogenital diseases | 45,951 | 39.4 (39.2 - 39.7) | 30,952 | 38.3 (38.0 - 38.7) | 26,060 | 37.8 (37.4 - 38.2) |
| °°° incontinence | 44,396 | 38.1 (37.8 - 38.4) | 29,607 | 36.7 (36.3 - 37.0) | 24,865 | 36.1 (35.7 -36.4) |
| °°° sexual disorders | 2,203 | 1.9 (1.8 - 2.0) | 1,841 | 2.3 (2.2 - 2.4) | 1,639 | 2.4 (2.3 - 2.5) |
| °°° urethral stricture | 215 | 0.2 (0.2 - 0.2) | 146 | 0.2 (0.2 - 0.2) | 118 | 0.2 (0.1 - 0.2) |
| °° sensory disorders | 26,224 | 22.5 (22.3 - 22.7) | 20,473 | 25.4 (25.1 - 25.7) | 17,931 | 26.0 (25.7 - 26.3) |
| °°° vestibular disorders | 11,377 | 9.8 (9.6 - 9.9) | 9,130 | 11.3 (11.1 -11.5) | 8,189 | 11.9 (11.6 - 12.1) |
| °°° hearing disorder | 18,003 | 15.5 (15.2 - 15.7) | 13,941 | 17.3 (17.0 - 17.5) | 12,035 | 17.5 (17.2 - 17.7) |
| °°° taste and smelling disorders | 220 | 0.2 (0.2 - 0.2) | 161 | 0.2 (0.2 - 0.2) | 138 | 0.2 (0.2 - 0.2) |
| °° impairment of nutrition | 17,835 | 15.3 (15.1 - 15.5) | 8,710 | 10.8 (10.6 - 11.0) | 6,705 | 9.7 (9.5- 9.9) |
| °° chronic pain | 24,833 | 21.3 (21.1 - 21.6) | 20,167 | 25.0 (24.7 - 25.3) | 19,499 | 28.3 (27.9 - 28.6) |
| °° multi-resistant infection | 15,792 | 13.6 (13.4 - 13.8) | 7,402 | 9.2 (9.0 - 9.4) | 5,419 | 7.9 (7.7 -8.1) |
| °° fatigue | 11,179 | 9.6 (9.4 - 9.8) | 6,529 | 8.1 (7.9 - 8.3) | 5,375 | 7.8 (7.6 - 8.0) |
| ° Incidence of new medical diagnoses, n, % of population at risk | | | | | | |
| °° respiratory dysfunction | 18,899 | 20.7 (20.5 - 21.0) | 9,022 | 14.7 (14.4 -15.0) | 7,704 | 14.1 (13.8 - 14.4) |
| °° cardiovascular diseases | 12,893 | 26.5 (26.1 - 26.9) | 4,228 | 14.0 (13.6 - 14.4) | 3,800 | 14.0 (13.6 - 14.4) |
| °°° coronary heart disease and myocardial infarction | 7,647 | 10.5 (10.3 - 10.7) | 3,161 | 6.4 (6.2 - 6.6) | 2,623 | 6.1 (5.9 - 6.3) |
| °°° cardiomyopathy | 1,992 | 1.8 (1.7 - 1.9) | 775 | 1.0 (0.9 - 1.1) | 653 | 1.0 (0.9 - 1.1) |
| °°° heart failure | 15,064 | 19.5 (19.2 - 19.8) | 5,967 | 11.8 (11.5 - 12.1) | 5,515 | 12.3 (12.0 - 12.6) |
| °°° cardiac arrhythmias | 12,389 | 15.4 (15.2 - 15.7) | 4,106 | 7.9 (7.7 - 8.1) | 3,580 | 7.8 (7.6 - 8.1) |
| °° cerebrovascular diseases | 5,927 | 5.9 (5.8 - 6.1) | 2,704 | 4.0 (3.8 - 4.1) | 2,295 | 3.4 (3.7 - 4.1) |
| °° renal diseases | 18,801 | 24.5 (24.2 - 24.8) | 6,710 | 14.4 (14.0 - 14.7) | 5,814 | 14.2 (13.8 - 14.5) |
| °° hepatic diseases | 120 | 0.1 (0.1 - 0.1) | 48 | 0.1 (0.0 - 0.1) | 30 | 0.0 (0.0 - 0.1) |
| °° metabolic diseases | 5,109 | 8.1 (7.9 - 8.3) | 2,060 | 4.9 (4.7 - 5.1) | 1,732 | 4.7 (4.5 - 5) |
| °°° diabetes mellitus | 4,859 | 7.6 (7.4 - 7.8) | 1,935 | 4.5 (4.3 - 4.7) | 1,638 | 4.4 (4.2 -2.6) |
| °°° other metabolic diseases | 597 | 0.5 (0.5 - 0.6) | 301 | 0.4 (0.3 - 0.4) | 259 | 0.4 (0.3 - 0.4) |
| °° anaemia | 15,937 | 17.3 (17.1 -17.6) | 5,474 | 9.2 (9.0 - 9.4) | 4,730 | 8.8 (8.6 - 9.1) |
| °° neuromuscular/ musculoskeletal diseases | 24,756 | 27.8 (27.5 - 28.1) | 8,562 | 16.1 (15.8 - 16.4) | 7,613 | 16.0 (15.6 - 16.3) |
| °°° ICUAW/CIP/CIM | 2,470 | 2.1 (2.0 - 2.2) | 273 | 0.3 (0.3 - 0.4) | 232 | 0.3 (0.3 - 0.4) |
| °°° dysphagia | 7,572 | 6.9 (6.7 - 7.0) | 2,625 | 3.5 (3.4 - 3.6) | 2,277 | 3.5 (3.4 - 3.7) |
| °°° voice disorders | 983 | 0.9 (0.8 - 0.9) | 587 | 0.7 (0.7 - 0.8) | 495 | 0.7 (0.7 - 0.8) |
| °°° contractures | 1,982 | 1.7 (1.7 - 1.8) | 1,009 | 1.3 (1.2 - 1.4) | 774 | 1.1 (1.1 - 1.2) |
| °°° immobility | 21,653 | 22.8 (22.5 - 35.1) | 7,990 | 13.8 (13.5 - 14.0) | 6,950 | 13.5 (13.3 - 13.8) |
| °° decubitus | 14,832 | 13.9 (13.7 - 14.1) | 4,460 | 6.4 (6.2 - 6.6) | 3,891 | 6.3 (6.1 - 6.5) |
| °° complications of the tracheostoma | 859 | 0.7 (0.7 - 0.8) | 182 | 0.2 (0.2 -0.3) | 97 | 0.1 (0.1 -0.2) |
| °°° tracheal stenoses | 362 | 0.3 (0.3 - 0.3) | 129 | 0.2 (0.1 - 0.2) | 68 | 0.1 (0.1 - 0.1) |
| °° urogenital diseases | 20,510 | 25.4 (25.1 - 15.7) | 6,688 | 13.6 (13.3 -13.9) | 5,728 | 13.0 (12.7 - 13.3) |
| °°° incontinence | 20,480 | 24.7 (24.4 - 25.0) | 6,498 | 12.8 (12.5 - 13.1) | 5,566 | 12.3 (12.0 - 12.6) |
| °°° sexual disorders | 614 | 0.5 (0.5 - 0.6) | 428 | 0.5 (0.5 - 0.6) | 359 | 0.5 (0.5 - 0.6) |
| °°° urethral stricture | 164 | 0.1 (0.1 - 0.2) | 83 | 0.1 (0.1 - 0.1) | 63 | 0.1 (0.1 - 0.1) |
| °° sensory disorders | 10,879 | 12.5 (12.3 - 12.7) | 6,234 | 10.6 (10.4 -10.9) | 5,370 | 10.6 (10.4 - 10.9) |
| °°° vestibular disorders | 6,627 | 6.4 (6.2 - 6.5) | 3,682 | 5.2 (5.0 - 5.4) | 3,117 | 5.1 (5.0 - 5.3) |
| °°° hearing disorder | 7,197 | 7.5 (7.4 - 7.7) | 4,455 | 6.8 (6.6 - 7.0) | 3,861 | 6.8 (6.6 - 7.1) |
| °°° taste and smelling disorders | 137 | 0.1 (0.1 - 0.1) | 80 | 0.1 (0.1 - 0.1) | 65 | 0.1 (0.1 - 0.1) |
| °° impairment of nutrition | 13,251 | 12.6 (12.4 - 12.8) | 4,853 | 6.8 (6.6 - 7.0) | 4,000 | 6.3 (6.1 -6.5) |
| °° chronic pain | 12,416 | 12.9 (12.7 - 13.1) | 6,586 | 10.5 (10.3 - 10.8) | 5,671 | 10.9 (10.7 - 11.2) |
| °° Multi-resistant infections | 13,189 | 12.0 (11.8 - 12.2) | 3,835 | 5.4 (5.2 - 5.6) | 3,077 | 4.8 (4.7 - 5.0) |
| °° fatigue | 8,925 | 8.2 (8.1 - 8.4) | 4,305 | 5.8 (5.7 - 6.0) | 3,604 | 5.6 (5.4 - 5.8) |
| **2. Psychological Diagnoses** | | | | | | |
| ° Prevalence of psychological diagnoses |  |  |  |  |  |  |
| °° PTSD | 431 | 0.4 (0.3 - 0.4) | 377 | 0.5 (0.4 - 0.5) | 368 | 0.5 (0.5 - 0.6) |
| °° Depression | 31,953 | 27.4 (27.2 - 27.7) | 24,550 | 30.4 (30.1 - 30.7) | 21,367 | 31.0 (30.6 - 31.3) |
| °° Anxiety | 8,192 | 7.0 (6.9 - 7.2) | 6,506 | 9.1 (7.9 - 8.2) | 5,653 | 8.2 (8.0 - 8.4) |
| °° Sleeping disorders | 17,058 | 14.6 (14.4 - 14.8) | 12,923 | 16 (15.8 - 16.3) | 11,180 | 16.2 (15.9 - 16.5) |
| °° Substance abuse | 15,142 | 13.0 (12.8 - 13.2) | 11,358 | 14.1 (13.8 - 14.3) | 9,876 | 14.3 (14.1 - 14.6) |
| ° Incidence of new psychological diagnoses, n, % of population at risk |  |  |  |  |  |  |
| °° PTSD | 211 | 0.2 (0.2 - 0.2) | 121 | 0.2 (0.1 - 0.2) | 109 | 0.2 (0.1 - 0.2) |
| °° Depression | 9,878 | 11.7 (11.5 - 11.9) | 4,276 | 7.7 (7.5 - 7.9) | 3,388 | 7.1 (6.9 - 7.4) |
| °° Anxiety | 3,550 | 3.3 (3.2 - 3.4) | 1,983 | 2.7 (2.6 - 2.8) | 1,585 | 2.5 (2.4 - 2.6) |
| °° Sleeping disorders | 6,833 | 6.9 (6.8 - 7.1) | 3,569 | 5.3 (5.2 - 5.5) | 2,880 | 5.0 (4.8 - 5.2) |
| °° Substance abuse | 4,103 | 4.1 (4.0 - 4.2) | 1,868 | 2.7 (2.6 - 2.8) | 1,635 | 2.8 (2.6 - 2.9) |

| **Table S3:** Prevalent and incident impairments in hospital survivors, 1-12, 13-24 and 25-36 months after sepsis | | | | | | |
| --- | --- | --- | --- | --- | --- | --- |
|  | **Follow-Up after index admission** | | | | | |
|  | **1-12 months** | | **13-24 months** | | **36 months** | |
| **Survivors, n** | **116507** | **% (95% CI)** | **80742** | **% (95% CI)** | **68940** | **% (95% CI)** |
| **1. Medical impairment** |  |  |  |  |  |  |
| - Prevalence of medical impairment, n, % of survivors | 102,018 | 87.6 (87.4 - 87.8) | 73,828 | 91.4 (91.2 - 91.6) | 62,896 | 91.2 (91 - 91.4) |
| - New onset of medical impairments, n, % of survivors | 82,629 | 70.9 (70.7 - 71.2) | 49,486 | 61.3 (61 - 61.6) | 37,885 | 55.0 (54.6 - 55.3) |
| - Number of new onset diseases, mean (SD); median (IQR) | 2.2 (1.4); 2 (2) | | 1.7 (1.0); 1 (2) | | 1.7 (1.0); 1 (1) | |
| - Prevalence of mechanical ventilation, n, % of sepsis survivors | 2,607 | 2.2 (2.2 - 2.3) | 1,841 | 2.3 (2.2 - 2.4) | 1,605 | 2.3 (2.2 - 2.4) |
| - Incidence of mechanical ventilation, n, % of population at risk | 1,890 | 1.6 (1.6 - 1.7) | 906 | 1.1 (1.1 - 1.2) | 751 | 1.1 (1 - 1.2) |
| - Prevalence of dialysis, n, % of sepsis survivors | 6,527 | 5.6 (5.5 - 5.7) | 4,195 | 5.2 (5 - 5.4) | 3,433 | 5 (4.8 - 5.1) |
| - Incidence of dialysis, n, % of population at risk | 3,144 | 2.8 (2.7 - 2.9) | 1,040 | 1.4 (1.3 - 1.4) | 789 | 1.2 (1.1 - 1.3) |
| **2. Psychological impairment** |  |  |  |  |  |  |
| - Prevalence of psychological impairment, n, % of survivors | 50,950 | 43.7 (43.4 - 44) | 38,598 | 47.8 (47.5 - 48.1) | 33,431 | 48.5 (48.1 - 48.9) |
| - New onset of psychological impairment, n, % of survivors | 20,840 | 17.9 (17.7 - 18.1) | 10,296 | 12.8 (12.5 - 13) | 8,429 | 12.2 (12 - 12.5) |
| - Number of new onset diseases, mean (SD); median (IQR) | 1.2 (0.5); 1 (0) | | 1.2 (0.4); 1 (0) | | 1.1 (0.4); 1 (0) | |
| **3. Cognitive impairment** |  |  |  |  |  |  |
| - Prevalence of cognitive impairment, n, % of survivors | 37,275 | 32 (31.7 - 32.3) | 25,352 | 31.4 (31.1 - 31.7) | 21,425 | 31.1 (30.7 - 31.4) |
| - Incidence of cognitive impairment , n, % of survivors at risk | 15,955 | 18.5 (18.2 - 18.7) | 5,383 | 9.8 (9.5 - 10) | 4,807 | 9.8 (9.6 - 10.1) |
| **4. Dependency on chronic care** |  |  |  |  |  |  |
| - Nursing home residence, % | 22,468 | 19.3 (19.1 - 19.5) | 15,149 | 18.8 (18.5 - 19.0) | 12,676 | 18.4 (18.1 - 18.7) |
| - Care level according to German care level system*, % | 58,209 | 50.0 (49.7 - 50.2) | 40,936 | 50.7 (50.4 - 51.0) | 35,577 | 51.6 (51.2 - 52.0) |
| - New nursing home residence, % survivors at risk | 12,485 | 12.0 (11.8 - 12.2) | 2,223 | 3.3 (3.2 - 3.5) | 1,950 | 3.4 (3.3 - 3.5) |
| - New care level according to German care level system*, % survivors at risk | 23,572 | 31.5 (31.1 - 31.8) | 3,784 | 9.2 (9.0 - 9.5) | 4,272 | 11.8 (11.5 - 12.1) |
| **5. Total health care costs** |  |  |  |  |  |  |
| Total health care costs [€]**, mean (SD); median (IQR) | 14,891 (24,737); 7,055 (14,958) | | 11,503 (20,788); 5,040 (10,904) | | 10,521 (19,146); 4,607 (9,803) | |
| IQR = Interquartile range; SD = Standard deviation  * eligibility for long-term care benefits in line with the German Social Code  **Total health care costs include cost for hospitalizations, outpatient consultations, medication and treatments (e.g. physical or occupational therapy) and rehabilitation | | | | | | |

| **Table S4:** Comparison of 1-12, 13-24 and 25-36 months outcomes and costs of non-severe sepsis/severe sepsis survivors | | | | | | |
| --- | --- | --- | --- | --- | --- | --- |
|  | **Follow- up** | | | | | |
|  | **1-12 months** | | **13-24 months** | | **25-36 months** | |
|  |  | **% (95% CI)** |  | **% (95% CI)** |  | **% (95% CI)** |
| **1. Medical impairment** | | | | | | |
| - Prevalence of medical impairment, n, % of survivors | | | | | | |
| Non-severe sepsis | 6,9311 | 88.1 (87.9 - 88.3) | 50,582 | 90.8 (90.5 - 91) | 43,157 | 90.6 (90.3 - 90.9) |
| Severe sepsis | 32,707 | 86.4 (86.1 - 86.8) | 23,246 | 92.9 (92.6 - 93.2) | 19,739 | 92.6 (92.3 - 93) |
| p-value | < 0.001 |  | < 0.001 |  | < 0.001 |  |
| - New onset of medical impairments, n, % of survivors | | | | | | |
| Non-severe sepsis | 55,094 | 70 (69.7 - 70.4) | 33,918 | 60.9 (60.5 - 61.3) | 26,142 | 54.9 (54.4 - 55.3) |
| Severe sepsis | 27,535 | 72.8 (72.3 - 73.2) | 15,568 | 62.2 (61.6 - 62.8) | 11,743 | 55.1 (54.4 - 55.8) |
| p-value | < 0.001 |  | < 0.001 |  | 0.616 |  |
| **2. Psychological impairment** | | | | | | |
| - Prevalence of psychological impairment, n, % of survivors | | | | | | |
| Non-severe sepsis | 34,278 | 43.6 (43.2 - 43.9) | 26,185 | 47 (46.6 - 47.4) | 22,684 | 47.6 (47.2 - 48.1) |
| Severe sepsis | 16,672 | 44.1 (43.6 - 44.6) | 12,413 | 49.6 (49 - 50.2) | 10,747 | 50.4 (49.8 - 51.1) |
| p-value | 0.119 |  | < 0.001 |  | < 0.001 |  |
| - New onset of psychological impairments, n, % of survivors | | | | | | |
| Non-severe sepsis | 13,665 | 17.4 (17.1 - 17.6) | 8,394 | 15.1 (14.8 - 15.4) | 5,715 | 12 (11.7 - 12.3) |
| Severe sepsis | 7,175 | 19 (18.6 - 19.4) | 3,911 | 15.6 (15.2 - 16.1) | 2,714 | 12.7 (12.3 - 13.2) |
| p-value | < 0.001 |  | 0.039 |  | 0.007 |  |
| **3. Cognitive impairment** | | | | | | |
| - Prevalence of cognitive impairment, n, % of survivors | | | | | | |
| Non-severe sepsis | 25,316 | 32.2 (31.9 - 32.5) | 17,627 | 31.6 (31.2 - 32) | 14,892 | 31.3 (30.9 - 31.7) |
| Severe sepsis | 11,959 | 31.6 (31.1 - 32.1) | 7,725 | 30.9 (30.3 - 31.5) | 6,533 | 30.7 (30 - 31.3) |
| p-value | 0.049 |  | 0.032 |  | 0.109 |  |
| - Incidence of cognitive impairment, n, % of population at risk | | | | | | |
| Non-severe sepsis | 10,278 | 17.8 (17.4 - 18.1) | 3,721 | 9.7 (9.4 - 10) | 3,271 | 9.7 (9.4 - 10) |
| Severe sepsis | 5,677 | 19.9 (19.5 - 20.4) | 1,662 | 9.8 (9.4 - 10.3) | 1,536 | 10.1 (9.6 - 10.6) |
| p-value | < 0.001 |  | 0.779 |  | 0.192 |  |
| **4.Co-occurence of cognitive/psychological and medical impairments** | | | | | | |
| - Prevalent impairments in two domains, n, % of survivors | | | | | | |
| Non-severe sepsis | 31,398 | 39.9 (39.6 - 40.3) | 23,401 | 42 (41.6 - 42.4) | 20,038 | 42.1 (41.6 - 42.5) |
| Severe sepsis | 15,095 | 39.9 (39.4 - 40.4) | 10,833 | 43.3 (42.7 - 43.9) | 9,256 | 43.4 (42.8 - 44.1) |
| p-value | 0.951 |  | 0.001 |  | 0.001 |  |
| - Prevalent impairments in all three domains, n, % of survivors | | | | | | |
| Non-severe sepsis | 13,279 | 16.9 (16.6 - 17.1) | 9,457 | 17 (16.7 - 17.3) | 8,105 | 17 (16.7 - 17.4) |
| Severe sepsis | 6,497 | 17.2 (16.8 - 17.6) | 4,375 | 17.5 (17 - 18) | 3,777 | 17.7 (17.2 - 18.2) |
| p-value | 0.221 |  | 0.075 |  | 0.024 |  |
| - New onset of impairments in two domains, n, % of survivors | | | | | | |
| Non-severe sepsis | 15,631 | 19.9 (19.6 - 20.2) | 8,315 | 14.9 (14.6 - 15.2) | 5,591 | 11.7 (11.5 - 12) |
| Severe sepsis | 8,333 | 22 (21.6 - 22.4) | 3,945 | 15.8 (15.3 - 16.2) | 2,677 | 12.6 (12.1 - 13) |
| p-value | < 0.001 |  | 0.002 |  | 0.002 |  |
| - New onset of impairments in all three domains, n, % of survivors | | | | | | |
| Non-severe sepsis | 2,711 | 3.4 (3.3 - 3.6) | 1,077 | 1.9 (1.8 - 2.1) | 636 | 1.3 (1.2 - 1.4) |
| Severe sepsis | 1,730 | 4.6 (4.4 - 4.8) | 481 | 1.9 (1.8 - 2.1) | 311 | 1.5 (1.3 - 1.6) |
| p-value | < 0.001 |  | 0.943 |  | 0.209 |  |
| **5. Dependence on chronic care** | | | | | | |
| - Nursing home residence, % | | | | | | |
| Non-severe sepsis | 15,013 | 19.1 (18.8 - 19.4) | 10,330 | 18.5 (18.2 - 18.9) | 8,654 | 18.2 (17.8 - 18.5) |
| Severe sepsis | 7,455 | 19.7 (19.3 - 20.1) | 4,819 | 19.3 (18.8 - 19.8) | 4,022 | 18.9 (18.4 - 19.4) |
| p-value | 0.013 |  | 0.015 |  | 0.029 |  |
| - Care level according to German care level system*, % | | | | | | |
| Non-severe sepsis | 39,206 | 49.8 (49.5 - 50.2) | 27,760 | 49.8 (49.4 - 50.2) | 24,181 | 50.8 (50.3 - 51.2) |
| Severe sepsis | 19,003 | 50.2 (49.7 - 50.7) | 13,176 | 52.7 (52 - 53.3) | 11,396 | 53.5 (52.8 - 54.1) |
| p-value | 0.225 |  | < 0.001 |  | < 0.001 |  |
| - Incident nursing home residence, % | | | | | | |
| Non-severe sepsis | 8,060 | 11.5 (11.3 - 11.7) | 1,573 | 3.4 (3.2 - 3.6) | 1,386 | 3.5 (3.3 - 3.7) |
| Severe sepsis | 4,425 | 13.1 (12.7 - 13.4) | 650 | 3.2 (3 - 3.4) | 564 | 3.2 (3 - 3.5) |
| p-value | < 0.001 |  | 0.148 |  | 0.096 |  |
| - New care level according to German care level system*, % | | | | | | |
| Non-severe sepsis | 14,931 | 29.9 (29.5 - 30.3) | 4,000 | 13.1 (12.8 - 13.5) | 3,019 | 11.9 (11.5 - 12.3) |
| Severe sepsis | 8,641 | 34.8 (34.2 - 35.3) | 1,645 | 13 (12.4 - 13.6) | 1,253 | 11.7 (11.1 - 12.3) |
| p-value | < 0.001 |  | 0.674 |  | 0.736 |  |
| **6. Health care costs** | | | | | | |
| - Total health care costs**, mean (SD) | | | | | | |
| Non-severe sepsis | 14,372 (24,289) | | 11,057 (20,579) | | 10,205 (19,339) | |
| Severe sepsis | 15,969 (25,610) | | 12,498 (21,213) | | 11,226 (18,687) | |
| p-value | <0.001 | | <0.001 | | <0.001 | |
| SD = Standard deviation  * eligibility for long-term care benefits in line with the German Social Code  ** Total health care costs include cost for hospitalizations, outpatient consultations, medication and treatments (e.g. physical or occupational therapy) and rehabilitation | | | | | | |

| **Table S5**: Comparison of 1-12, 13-24 and 25-36 months outcomes and costs of ICU-treated/non-ICU-treated sepsis survivors | | | | | | |
| --- | --- | --- | --- | --- | --- | --- |
|  | **Follow-up** | | | | | |
|  | **1-12 months** | | **13-24 months** | | **25-36 months** | |
|  | **n** | **% (95% CI)** | **n** | **% (95% CI)** | **n** | **% (95% CI)** |
| **1. Medical diagnosis** | | | | | | |
| - Prevalence of medical diagnosis, n, % of survivors | | | | | | |
| Non-ICU-treated sepsis | 73,907 | 87.7 (87.5 - 87.9) | 53,558 | 91.1 (90.9 - 91.4) | 45,560 | 90.9 (90.6 - 91.1) |
| ICU-treated sepsis | 28,111 | 87.2 (86.8 - 87.6) | 20,270 | 92.2 (91.9 - 92.6) | 17,336 | 92.2 (91.8 - 92.6) |
| p-value | 0.019 |  | < 0.001 |  | < 0.001 |  |
| - New onset of medical diagnosis, n, % of survivors | | | | | | |
| Non-ICU-treated sepsis | 58,369 | 69.3 (69 - 69.6) | 35,984 | 61.2 (60.8 - 61.6) | 27,688 | 55.2 (54.8 - 55.7) |
| ICU-treated sepsis | 24,260 | 75.3 (74.8 - 75.7) | 13,502 | 61.4 (60.8 - 62.1) | 10,197 | 54.2 (53.5 - 55) |
| p-value | < 0.001 |  | 0.624 |  | 0.022 |  |
| **2. Psychological diagnosis** | | | | | | |
| - Prevalence of psychological diagnosis, n, % of survivors | | | | | | |
| Non-ICU-treated sepsis | 35,915 | 42.6 (42.3 - 43.0) | 27,240 | 46.4 (46.0 - 46.8) | 23,596 | 47.1 (46.6 - 47.5) |
| ICU-treated sepsis | 15,035 | 46.6 (46.1 - 47.2) | 11,358 | 51.7 (51.0 - 52.3) | 9,835 | 52.3 (51.6 - 53.0) |
| p-value | < 0.001 |  | < 0.001 |  | < 0.001 |  |
| - New onset of psychological diagnosis, n, % of survivors | | | | | | |
| Non-ICU-treated sepsis | 13,937 | 16.5 (16.3 - 16.8) | 7,309 | 12.4 (12.2 - 12.7) | 5,989 | 11.9 (11.7 - 12.2) |
| ICU-treated sepsis | 6,903 | 21.4 (21.0 - 21.9) | 2,987 | 13.6 (13.1 - 14.0) | 2,440 | 13.0 (12.5 - 13.5) |
| p-value | < 0.001 |  | < 0.001 |  | < 0.001 |  |
| **3. Cognitive diagnosis** | | | | | | |
| - Prevalence of cognitive diagnosis, n, % of survivors | | | | | | |
| Non-ICU-treated sepsis | 28,011 | 33.2 (32.9 - 33.6) | 19,329 | 32.9 (32.5 - 33.3) | 16,203 | 32.3 (31.9 - 32.7) |
| ICU-treated sepsis | 9,264 | 28.7 (28.2 - 29.2) | 6,023 | 27.4 (26.8 - 28.0) | 5,222 | 27.8 (27.1 - 28.4) |
| p-value | < 0.001 |  | < 0.001 |  | < 0.001 |  |
| - Incidence of cognitive diagnosis , n, % of population at risk | | | | | | |
| Non-ICU-treated sepsis | 10,728 | 17.8 (17.4 - 18.1) | 3,938 | 9.9 (9.7 - 10.2) | 3,451 | 9.9 (9.6 - 10.2) |
| ICU-treated sepsis | 5,227 | 20.2 (19.7 - 20.7) | 1,445 | 9.3 (8.8 - 9.8) | 1,356 | 9.7 (9.3 - 10.2) |
| p-value | < 0.001 |  | 0.019 |  | 664 |  |
| **4.Co-occurence of cognitive/psychological and medical diagnoses** | | | | | | |
| - Prevalent diagnoses in two domains, n, % of survivors | | | | | | |
| Non-ICU-treated sepsis | 33,300 | 39.5 (39.2 - 39.8) | 24,539 | 41.8 (41.4 - 42.2) | 20,969 | 41.8 (41.4 - 42.3) |
| ICU-treated sepsis | 13,193 | 40.9 (40.4 - 41.5) | 9,695 | 44.1 (43.5 - 44.8) | 8,325 | 44.3 (43.6 - 45.0) |
| p-value | < 0.001 |  | < 0.001 |  | < 0.001 |  |
| - Prevalent diagnoses in all three domains, n, % of survivors | | | | | | |
| Non-ICU-treated sepsis | 14,509 | 17.2 (17.0 - 17.5) | 10,276 | 12.2 (12.0 - 12.4) | 8,766 | 10.4 (10.2 - 10.6) |
| ICU-treated sepsis | 5,267 | 16.3 (15.9 - 16.7) | 3,556 | 11.0 (10.7 - 11.4) | 3,116 | 9.7 (9.3 - 10.0) |
| p-value | < 0.001 |  | < 0.001 |  | 0.005 |  |
| - New onset of diagnoses in two domains, n, % of survivors | | | | | | |
| Non-ICU-treated sepsis | 16,162 | 19.2 (18.9 - 19.4) | 8,765 | 14.9 (14.6 - 15.2) | 5,922 | 11.8 (11.5 - 12.1) |
| ICU-treated sepsis | 7,802 | 24.2 (23.7 - 24.7) | 3,495 | 15.9 (15.4 - 16.4) | 2,346 | 12.5 (12 - 13) |
| p-value | < 0.001 |  | 0.001 |  | 0.017 |  |
| - New onset of diagnoses in all three domains, n, % of survivors | | | | | | |
| Non-ICU-treated sepsis | 2,763 | 3.3 (3.2 - 3.4) | 1,096 | 1.9 (1.8 - 2) | 673 | 1.3 (1.2 - 1.4) |
| ICU-treated sepsis | 1,678 | 5.2 (5 - 5.5) | 462 | 2.1 (1.9 - 2.3) | 274 | 1.5 (1.3 - 1.6) |
| p-value | < 0.001 |  | 0.032 |  | 0.262 |  |
| **5. Dependence on nursing care** | | | | | | |
| - Nursing home residence, % | | | | | | |
| Non-ICU-treated sepsis | 16,840 | 20.0 (19.7 - 20.3) | 11,496 | 19.6 (19.2 - 19.9) | 9,617 | 19.2 (18.8 - 19.5) |
| ICU-treated sepsis | 5,628 | 17.5 (17.0 - 17.9) | 3,653 | 16.6 (16.1 - 17.1) | 3,059 | 16.3 (15.8 - 16.8) |
| p-value | < 0.001 |  | < 0.001 |  | < 0.001 |  |
| - Care level according to German care level system*, % | | | | | | |
| Non-ICU-treated sepsis | 42,305 | 50.2 (49.9 - 50.5) | 29,704 | 50.5 (50.1 - 51.0) | 25,780 | 51.4 (51.0 - 51.9) |
| ICU-treated sepsis | 15,904 | 49.3 (48.8 - 49.9) | 11,232 | 51.1 (50.4 - 51.8) | 9,797 | 52.1 (51.4 - 52.8) |
| p-value | 0.008 |  | < 0.001 |  | < 0.001 |  |
| - Incident home residence, % | | | | | | |
| Non-ICU-treated sepsis | 8,535 | 11.5 (11.3 - 11.8) | 1,703 | 3.5 (3.4 - 3.7) | 1,502 | 3.6 (3.4 - 3.8) |
| ICU-treated sepsis | 3,950 | 13.2 (12.8 - 13.5) | 520 | 2.8 (2.6 - 3.1) | 448 | 2.8 (2.6 – 3.1) |
| p-value | < 0.001 |  | < 0.001 |  | < 0.001 |  |
| - Incident care level according to German care level system*, % | | | | | | |
| Non-ICU-treated sepsis | 15,041 | 29.1 (28.7 - 29.5) | 2,797 | 9.3 (9.0 - 9.6) | 3,190 | 12.0 (11.7 - 12.4) |
| ICU-treated sepsis | 8,531 | 36.8 (36.2 - 37.5) | 987 | 9.1 (8.6 – 9.7) | 1,082 | 11.2 (10.6 – 11.8) |
| p-value | < 0.001 |  | 0.577 |  | 0.023 |  |
| **6. Health care costs** | | | | | | |
| - Total health care costs**, mean (SD) | | | | | | |
| Non-ICU-treated sepsis | 13,682 (23,214) | | 10,831 (19,764) | | 10,020 (17,905) | |
| ICU-treated sepsis | 18,051 (28,090) | | 13,300 (23,213) | | 11,857 (22,062) | |
| p-value | < 0.001 | | < 0.001 | | < 0.001 | |
| SD = Standard deviation; * eligibility for long-term care benefits in line with the German Social Code; **Total health care costs include cost for hospitalizations, outpatient consultations, medication and treatments (e.g. physical or occupational therapy) and rehabilition | | | | | | |

| **Table S6**: 1-12, 13-24 and 25-36 months outcomes and costs of patients without pre-existing impairments | | | | | | |
| --- | --- | --- | --- | --- | --- | --- |
|  | **Follow-up** | | | | | |
|  | **1-12 months** | | **13-24 months** | | **25-36 months** | |
|  | **n** | **% (95% CI)** | **n** | **% (95% CI)** | **n** | **% (95% CI)** |
| **Survivors, n** | 8,622 |  | 7,314 |  | 6,869 |  |
| **1. Medical diagnosis** |  |  |  |  |  |  |
| - Prevalence of medical diagnosis, n, % of survivors | 5,472 | 63.5 (62.4 - 64.5) | 4,382 | 59.9 (58.8 - 61) | 4,236 | 61.7 (60.5 - 62.8) |
| - New onset of medical diagnosiss, n, % of survivors | 5,472 | 63.5 (62.4 - 64.5) | 2,942 | 40.2 (39.1 - 41.4) | 2,437 | 35.5 (34.4 - 36.6) |
| **2. Psychological diagnosis** |  |  |  |  |  |  |
| - Prevalence of psychological diagnosis, n, % of survivors | 2,157 | 25 (24.1 - 25.9) | 1,820 | 24.9 (23.9 - 25.9) | 1,790 | 26.1 (25 - 27.1) |
| - New onset of psychological diagnosis, n, % of survivors | 2,157 | 25 (24.1 - 25.9) | 880 | 12 (11.3 - 12.8) | 685 | 10 (9.3 - 10.7) |
| **3. Cognitive diagnosis** | | | | | | |
| - Prevalence of cognitive diagnosis, n, % of survivors | 1,106 | 12.8 (12.1 - 13.6) | 834 | 11.4 (10.7 - 12.2) | 818 | 11.9 (11.2 - 12.7) |
| - Incidence of cognitive diagnosis, n, % of survivors at risk | 1,106 | 12.8 (12.1 - 13.6) | 327 | 4.5 (4.0 - 5.0) | 244 | 3.6 (3.1 - 4.0) |
| **4.Co-occurence of cognitive/psychological and medical diagnosis** | | | | | | |
| - Prevalent diagnoses in two domains, n, % of survivors | 1,905 | 22.1 (21.2 - 23) | 1,543 | 21.1 (20.2 - 22) | 1,501 | 21.9 (20.9 - 22.8) |
| - Prevalent diagnoses in all three domains, n, % of survivors | 462 | 5.4 (4.9 - 5.9) | 320 | 4.4 (3.9 - 4.9) | 331 | 4.8 (4.3 - 5.4) |
| - New onset of diagnoses in two domains, n, % of survivors | 1,905 | 22.1 (21.2 - 23) | 679 | 9.3 (8.6 - 10) | 526 | 7.7 (7.1 - 8.3) |
| - New onset of diagnoses in all three domains, n, % of survivors | 462 | 5.4 (4.9 - 5.9) | 65 | 0.9 (0.7 - 1.1) | 41 | 0.6 (0.4 - 0.8) |
| **5. Dependency on chronic care** | | | | | | |
| - Nursing home residence, % | 663 | 7.7 (7.1 - 8.3) | 466 | 6.4 (5.8 - 7) | 459 | 6.7 (6.1 - 7.3) |
| - Care level according to German care level system*, % | 1,979 | 23 (22.1 - 23.9) | 1,498 | 20.5 (19.6 - 21.4) | 1,489 | 21.7 (20.7 - 22.7) |
| - New nursing home residence, % survivors at risk | 615 | 7.2 (6.7 - 7.8) | 91 | 1.3 (1.1 - 1.6) | 88 | 1.4 (1.1 - 1.7) |
| - New care level according to German care level system*, % survivors at risk | 1,582 | 19.3 (18.5 - 20.2) | 216 | 3.7 (3.2 - 4.2) | 283 | 5.1 (4.5 - 5.7) |
| **6. Total health care costs** | | | | | | |
| Total health care costs [€]**, mean (SD); median (IQR) | 12,583 (25,932); 3,716 (11,884) | | 7,004 (20,972); 1,486 (5,260) | | 5,621 (14,215); 1,312 (4,100) | |
| IQR=interquartile range; SD=standard deviation; * eligibility for long-term care benefits in line with the German Social Code; ** Total health care costs include cost for hospitalizations, outpatient consultations, medication and treatments (e.g. physical or occupational therapy) and rehabilitation | | | | | | |

| **Table S7: Total health care costs hospital survivors, 1-12, 13-24 and 25-36 months after sepsis** | | | |
| --- | --- | --- | --- |
|  | **Sepsis** | **Severe Sepsis** | **Non-Severe Sepsis** |
| **n** | 159,684 | 69,956 | 89,728 |
| 12 months prior to index, mean (SD). median (IQR) | 13,074 (19,690); 6,327 (13,864) | 13,192 (20,233); 6,495 (13,912) | 12,982 (19,255); 6,194 (13,826) |
| Index hospitalization, mean (SD). median (IQR) | 15,361 (28,731); 5,849 (10,275) | 22,496 (36,840); 9,281 (22,924) | 9,797 (18,446); 4,410 (5,205) |
| 12 months after index, mean (SD). median (IQR) | 10,865 (22,140); 3,394 (12,339) | 8,639 (20,446); 112 (8,939) | 12,601 (23,228); 5,217 (13,459) |
| 24 months after index, mean (SD). median (IQR) | 5,817 (15,861); 43 (5,138) | 4,470 (14,030); 0 (2,699) | 6,866 (17,081); 1,380 (6,736) |
| 36 months after index, mean (SD). median (IQR) | 4,542 (13,616); 0 (3,487) | 3,420 (11,536); 0 (1,383) | 5,417 (14,982); 441 (4881) |
| Total costs of index hospitalization and 0-36 months follow up* | 36,585 (50,368); 21,029 (35,394) | 39,025 (53,209); 22,259 (42,029) | 34,682 (47,951); 20,283 (31,204) |
| IQR = Interquartile range; SD = Standard deviation; *Total health care costs include cost for hospitalizations, outpatient consultations, medication and treatments (e.g. physical or occupational therapy) and rehabilitation | | | |

**Fig. S1:** Co-occurrence of diagnoses and mortality in patients 1-12 months after discharge from the index hospitalization according to pre-existing diagnoses

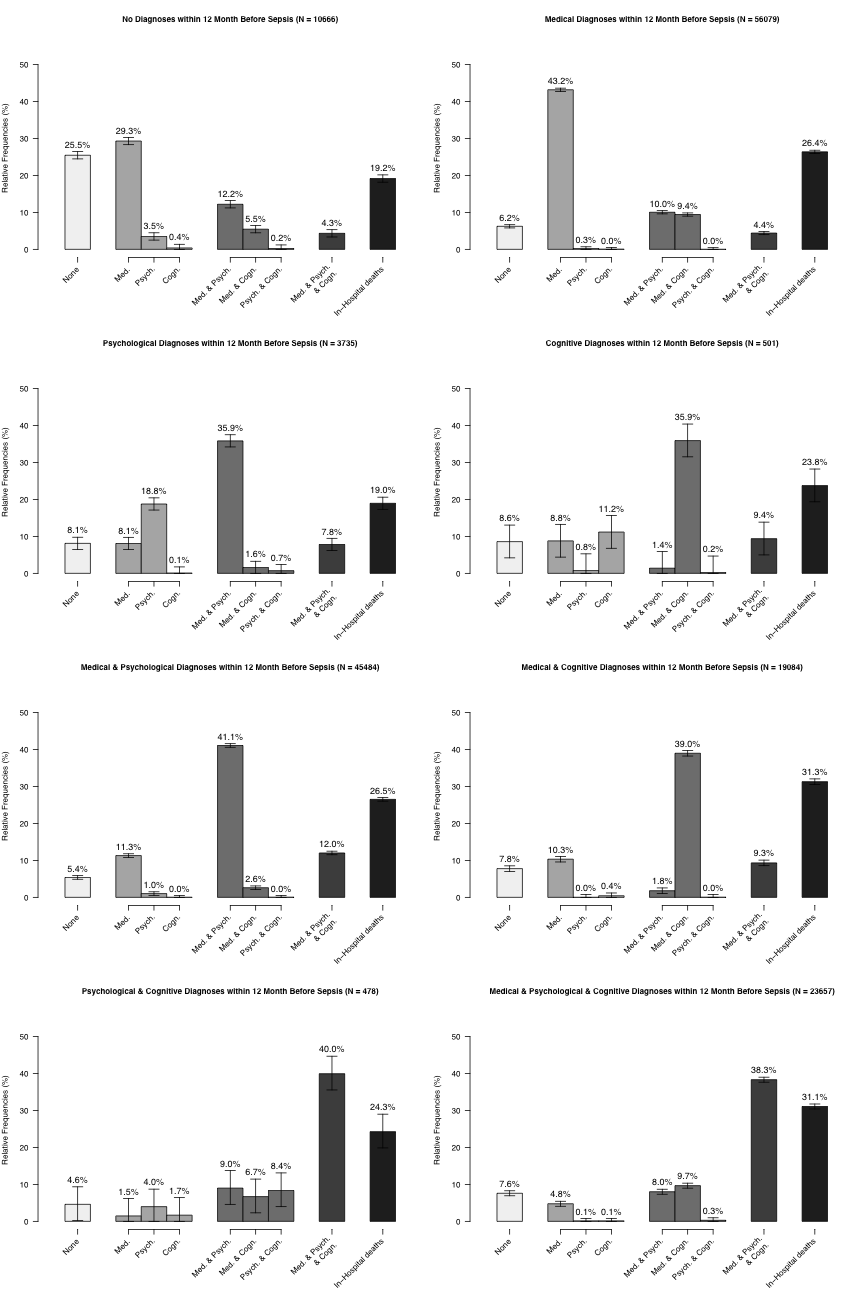

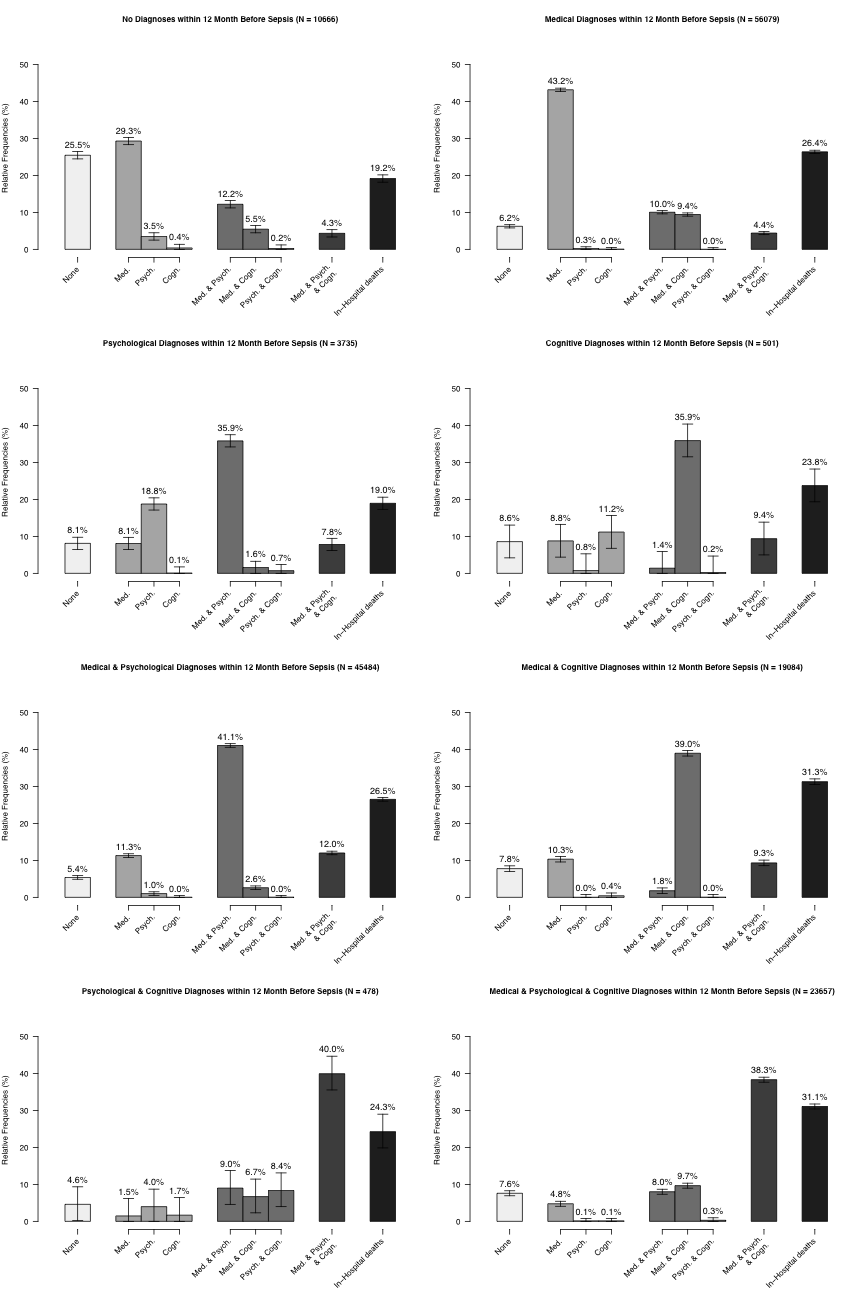

**Fig. S3:** Hazard functions for death for a) all sepsis patients, b) severe and non-severe sepsis patients, c) ICU- and non-ICU-treated sepsis patients, d) sepsis patients according to pre-existing impairments, and e) age groups

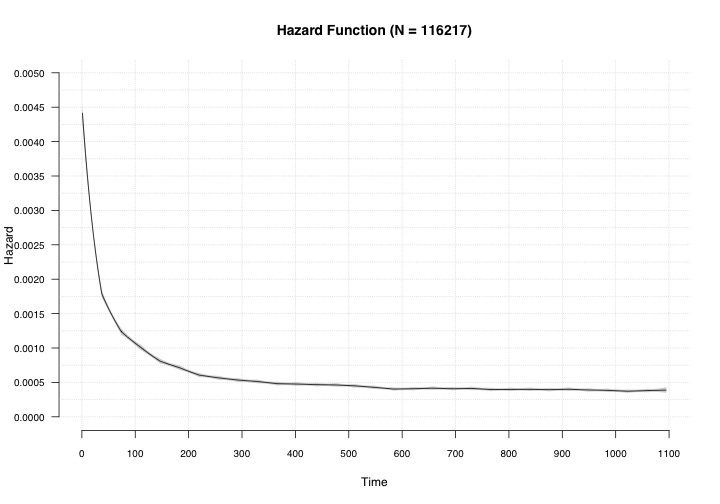

**(a)**

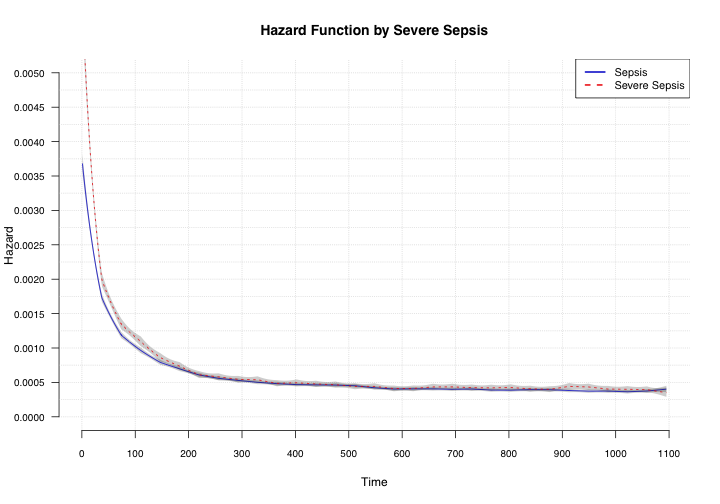

**(b)**

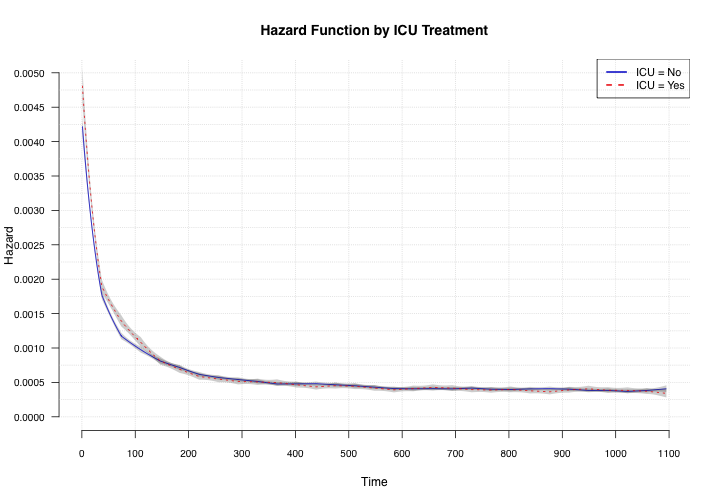

**(c)**

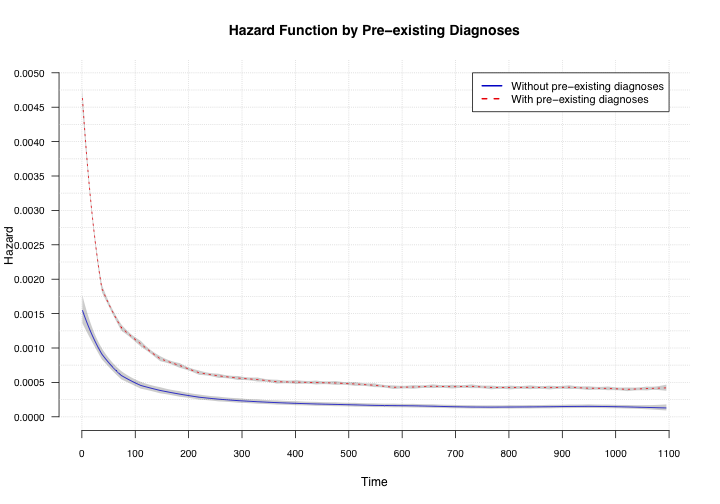

**(d)**

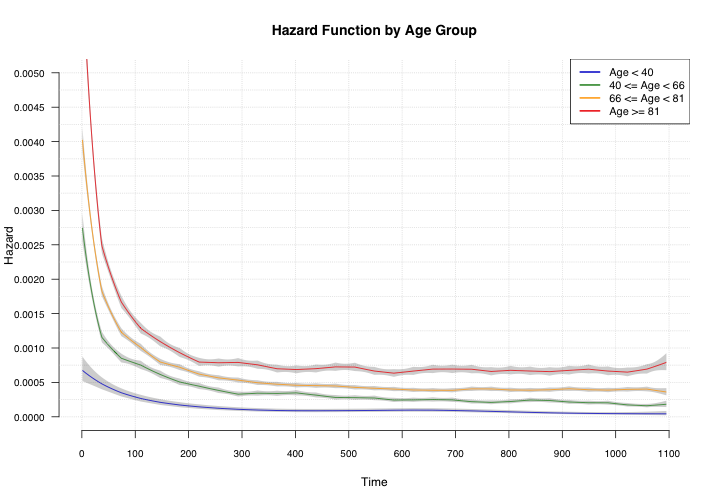

**(e)**
